# Effects of polygenic risk score communication on health outcomes: systematic review and meta-analysis

**DOI:** 10.1101/2025.11.04.25339466

**Authors:** Luigi Russo, Nicolò Lentini, Sara Farina, Antonio Cristiano, Andrea Adduci, Alessio Perilli, Christian Cao, Roberta Pastorino, Stefania Boccia, John PA Ioannidis

## Abstract

**Objective:** The purpose of this systematic review and meta-analysis is to summarize evidence from all RCTs to-date on the efficacy of polygenic risk score (PRS) communication in changing health outcomes.

**Design:** Systematic review and meta-analysis.

**Data sources:** Cochrane Central Register of Controlled Trials (CENTRAL) and Pubmed, from inception to March 2025.

**Study selection:** Randomized controlled trials comparing disclosure versus non-disclosure of PRS results. Intervention included comunication of risk information from ≥ 2 single nucleotide polymorphism (SNPs).

**Methods:** Pairs of reviewers conducted screening, extracted data and assessed risk of bias. Meta-analyses were conducted using inverse variance-weighting with fixed and random effects models. Otto-SR was used to verify screening and data extraction. Risk of bias was assessed with the Cochrane Risk of Bias 2 (RoB-2) tool. All measurable health-related outcomes were considered eligible.

**Results:** Of 7,830 articles retrieved, 27 RCTs were eligible. PRSs mainly predicted risks for cancer (n=9 RCTs), cardiovascular diseases (n=8), and diabetes (n=6). 21 RCTs targeted primarily healthy populations, 3 at-risk populations and 3 individuals who had already developed a disease and PRS predicted complications. 15/26 RCTs concluded in their abstracts with favorable claims about the PRS, with only 5/15 justifying it with any statistically significant results. 9/26 RCTs had high risk of bias. Meta-analysis revealed no statistically significant effects on any measured outcome, among 22 outcomes tested in 2 or more trials. Standardized mean differences (SMDs) (95% CI) for dietary outcomes were −0.11 (−0.23; 0.01) for daily energy intake, 0.08 (−0.15; 0.31) for daily fat intake and −0.11 (−0.28; 0.06) for alcohol consumption. For physical activity, SMD was −0.01 (−0.13; 0.11). Relative risks were 1.12 (0.77; 1.61) for screening attendance, 1.50 (0.98; 2.29) for statin use, and 0.95 (0.32; 2.79) for disease incidence. For psychological outcomes, SMDs were −0.02 (−0.13; 0.08) for anxiety, −0.06 (−0.23; 0.10) for worry, −0.10 (−0.40-0.19) for perceived risk, and −0.05 (−0.23; 0.13) for depression. For clinical outcomes, mean differences were −2.01 (−8.27; 4.26) for total cholesterol, −3.64 (−7.88; 0.60) for LDL cholesterol, −0.21 (−2.65; 2.23) for HDL cholesterol, −1.88 (−4.17;0.42) for diastolic blood pressure, −1.26 (−4.44; 1.92) for systolic blood pressure, −0.12 (−0.64; 0.39) for BMI and −0.33 (−0.87; 0.20) for weight. 69 outcomes had been reported in only a single trial (18/69 primary ones), and of those 2 had statistically significant results at p<0.05 as primary outcomes and 3 as secondary outcomes.

**Conclusions:** Overall, despite frequent promising claims, the disclosure of PRS typically did not lead to meaningful changes in behavioral, psychological or clinical outcomes.

**Systematic review registration:** OpenScience Framework: doi.org/10.17605/OSF.IO/28V6J

**Summary box:** *Section 1: What is already known on this topic:*

- Polygenic risk scores (PRSs) represents one of the most promising approaches of personalized medicine where communication of genetic information is expected to improve behavior and health outcomes.
- Previous reviews have evaluated the communication of genetic risk but none have evaluated yet the effect of specifically PRS.
- Since PRSs implementation in clinical practice is contemplated, rigorous evaluation of their impact is valuable for further implementation.

*Section 2: What this study adds:*

- PRS communication effects were close to null for a large set of outcomes, demonstrating no improvement in preventive behaviours, including screening adherence or clinical measures.
- High heterogeneity in RCTs in the field is present with small sample sizes, short follow-up periods, and many self-reported outcomes.
- A gap is evident between the theoretical promise of PRS-guided prevention and its lack of documented real-world effectiveness.

## INTRODUCTION

Personalized medicine using genetic and genomic information has long promised a revolution in disease prevention, diagnosis and treatment (1,2). While major advancements have occurred for genomic-based testing on cancer tissue for guiding treatment (3,4) and for genetic testing applications for rare disease diagnosis (5,6), genomic-testing applications for disease prevention of common, multigenetic diseases remain limited. While guidelines using genetic testing to prevent and risk stratify hereditary cancers and cardiovascular diseases have existed for decades (7,8), the application of sophisticated algorithms that combine the effects of low- to moderate-penetrance genetic variants is still in its infancy, largely due to the lack of robust evidence on clinical utility (9,10).

Thousands of studies have examined the relationship between genetic variants and specific traits, such as diseases and certain conditions. Genome-wide assessments have detected large numbers of single nucleotide polymorphisms (SNPs) that occur more frequently in affected individuals, providing insights into genetic risk factors. Weighted sums, calculated from the identified SNPs, are usually referred to as Polygenic Risk Scores (PRSs, aka, genetic risk scores or polygenic scores) (11,12). These scores, which quantify an individual’s genetic predisposition to a disease, are often presented as promising tools for personalized medicine, given their potential in disease risk prediction (13,14).

PRSs could enable the identification of high-risk individuals, who, in principle, could be targeted through personalized preventive strategies. Since PRS can be constructed using a range of common genetic variants—from a large number, as in a genome-wide PRS; to only a few SNPs, known as restricted PRS—their classification varies widely in the current literature (15,16).

Several systematic reviews and meta-analyses have previously synthesized data from RCTs comparing groups receiving genetic risk information to those who did not (17–21). These studies have examined various outcomes, including smoking cessation, healthier dietary habits, increased physical activity, and changes in cholesterol levels or other disease risk factors. However, findings have been inconclusive and each review only included 10 unique RCTs that have used PRS genetic information that extends beyond a single SNP or non-pathogenetic variant.

In recent years, several RCTs have explored the impact of PRS-based communication on diverse outcomes, ranging from lifestyle changes and adherence to screening protocols. Moreover, more comprehensive PRSs have been used in the more recent trials and a wider range of PRSs has been assessed.

The purpose of this systematic review and meta-analysis is to summarize evidence from all RCTs to-date on the efficacy of PRS communication in changing health behaviors, therapy adherence, psychological outcomes, and clinical outcomes.

## METHODS

We previously registered the protocol for this systematic review and meta-analysis on OpenScienceFramework (22). This systematic review is reported following the Preferred Reporting Items for Systematic Reviews and Meta-Analyses (PRISMA) Checklist (23).

### Definitions

The National Cancer Institute defines PRSs as an assessment of the risk for a specific condition based on the cumulative influence of multiple genetic variants (24–27). These scores are calculated by aggregating and quantifying the effects of common genetic variants, typically SNPs, i.e. variations at a single base pair within the genome, classified as minor alleles with a frequency greater than 1%. Individually, these variants have small effects, but when combined, they contribute to the overall risk score. The resulting PRS generally follows a normal distribution within the population, where higher scores indicate a greater risk of developing the condition. SNPs included in a PRS can range widely depending on the methodology used and the variants selection, ranging from only a few to thousands.

### Search strategy

We conducted a preliminary search of PubMed and Scopus to identify relevant keywords and previous systematic reviews on the same theme. Then, we used the terms identified during this process to build our search strategy. Firstly, we attempted to update previous systematic reviews by reusing their search strategies. However, re-running the search strings from Horne et al. (2017), from 2017 to the present, yielded over 70,000 results. Similarly, the search string used in Hollands et al. (2016), restricted to records from 2016 onward, returned 21,324 results only in Medline. Due to this explosion of literature, previous search strategies were not deemed efficient to apply. As such we extracted all the unique trials contained in the systematic reviews previously identified during the preliminary search (17–21) and we built our search string using the following terms:

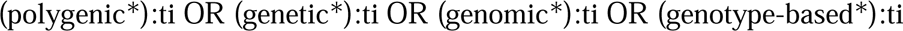

This string, when applied to Pubmed with the filter for ‘Clinical trials’ and ‘Randomized Controlled Trials’, was able to identify 35 out of 37 trials included in previous systematic reviews and perusal of the references of these 35 would yield also the other 2 missing trials. The string also captured all 10 unique trials that had been included in at least one previous systematic review dealing with PRSs that would be eligible for our protocol. This string was then applied to identify articles on Pubmed, with the RCTs and CTs filter, and on Cochrane Central Register of Controlled Trials (CENTRAL) without restrictions on the publication year. Furthermore, we searched on ClinicalTrials.gov with the following terms combination “Polygenic Risk Score” OR “polygenic score” OR “genetic risk score” OR “genetic score”.

We retrieved articles up to the 1^st^ of March 2025. We also examined the references of the retrieved eligible articles for any additional relevant RCTs, and we searched for each one of the included RCT the registration on clinicaltrials.gov for any available study results.

### Study eligibility and selection

We evaluated each article against all the following eligibility criteria: (1) RCTs involving humans; (2) articles published or accepted for publication (i.e., in press) in peer–reviewed journals, or with available results on ClinicalTrial.Gov, without time restriction in any language; (3) RCTs comparing the disclosure of inherited genetic risk obtained through the PRS for any phenotype (intervention group), with scenarios where no genetic information is provided (comparator group). Quasi-randomized clinical trials, cluster randomized clinical trials, and all non-randomized studies were excluded. Studies focusing on high-penetrant genes or monogenic disorders, such as *BRCA1/2* mutations, were also excluded.

We included all studies that defined PRS as above, even if it was not explicitly referred to as “PRS” but, for example, it was reported as “genetic risk score”, genetic score”, or “polygenic score”. At least 2 SNPs were needed to classify a score as PRS.

### Study selection

We uploaded the identified articles to the Rayyan platform, a software for systematic reviews. After deduplication, four reviewers (L.R., A.A., S.F., A.C.) conducted title and abstract screening and then reviewed the full text, with each record assessed by two independent reviewers. Any discrepancies were addressed by consensus or by involving a third reviewer. Finally, we included the articles that met the eligibility criteria in the systematic review. For included studies that did not report results (e.g. abstracts or protocols), we contacted study authors and principal investigors requesting data, if available.

### Data extraction

Three reviewers (L.R., A.C., N.L.) independently performed data extraction. They extracted the following data from each included study: title, first author, doi, journal of publication, year of publication, country, eligibility criteria, demographic information, study design, number of arms, type of information for each arm, number of participants in each arm, possible phenotypes predicted by the PRS, number of SNPs, risk of developing those phenotypes, target population of the intervention (i.e., healthy, at risk, etc.), and the outcomes of the study.

For each study, we extracted the following outcomes: behavioral changes (i.e., dietary behavior, physical activity, smoking, alcohol consumption, attendance for medical visits like check-up, cancer screenings, follow-up visits, undergoing risk-reducing surgery, or other behaviors aimed at reducing disease risk), adherence to therapy, incidence of the disease for which the genetic information is provided, quality of life, psychological impact (e.g. depression, anxiety, emotional distress, worry, perceived efficacy, perceived risk, or other measured psychological change), incidence of other diseases (other than the disease linked to the genetic risk score), any other measurable health outcome or result of the trial.

For each outcome, we extracted the measurement, and the method used. When an outcome is measured subjectively, we refer to self-reported data from the patient. In contrast, for an objective measurement we refer to data obtained through tests, analyses, or medical evaluations.

Although not pre-specified in the protocol, we conducted a post-hoc data extraction from the registry in which each included RCT was registered (e.g., ClinicalTrials.gov, UMIN-CTR, ISRCTN). For each trial, we collected information on the primary and secondary outcomes reported and any available results. In addition, from each article we extracted the conclusions stated in the abstract and assessed whether, and how, the results supported those conclusions.

### Screening and data extraction validation with Otto-SR

As an additional validation step, we used an artificial intelligence platform, otto-SR (28), to validate the retrieval of eligible articles and the correctness of data extraction. Otto-SR is an LLM-based tool for automated screening and data extraction that was developed after our protocol had been posted.

For the screening phase, Otto-SR conducted screening of the PubMed and CENTRAL results but did not screen the ClinicalTrials.gov search results. The outputs generated by Otto-SR were compared with those obtained by human reviewers, and concordance was calculated as the proportion of full-text articles correctly identified by both approaches over the total number of studies included in the review. Discrepancies were classified as inclusion errors (articles included by Otto-SR but not by humans) or exclusion errors (articles missed by Otto-SR but included by humans). For the data extraction phase, the information independently extracted by Otto-SR and the human reviewer was compared across all predefined data fields. Concordance was defined as identical extraction between the two approaches. Discrepancies were categorized as extraction errors by Otto-SR or by the human reviewer, and overall concordance rates and error proportions were calculated for each dataset.

### Risk of bias assessment

Three authors (L.R., S.F., A.P.) assessed the risk of bias of the included studies using the Revised Cochrane Risk-of-Bias Tool for Randomized Trials (RoB 2) (29), independently from each other, with each study assessed twice.

### Statistical analysis

We reported the general characteristics of the studies. For the quantitative analyses of study results, for continuous outcomes we reported outcomes as means and standard deviations. We conducted the meta-analysis for studies that reported outcome measures either at both the start and end of the trial, or the difference in value. Whenever studies only provided the final value, meta-analysis was conducted on the final difference between arms. To compare the control group (participants without disclosure of inherited polygenic risk score) and the PRS group (participants with disclosure of PRS), we used the mean difference (MD) for continuous outcomes measured using the same method and unit across studies. When necessary, units were harmonized using appropriate conversion factors (e.g., converting LDL, HDL, and total cholesterol from mmol/L to mg/dL, and weight from pounds to kilograms). When outcomes were assessed using different measurement methods or scales across studies that could not be similarly converted to each other, we used the standardized mean difference (SMD) calculated using Hedges’ method (30). For binary outcomes, we used the relative risk (RR) as the effect size, comparing groups at the end of the study. All effect sizes were reported with 95% confidence intervals (CIs).

We additionally conducted two sensitivity analyses: one meta-analysis on final values including only studies that reported both baseline and final values, and another meta-analysis on final values including only studies that reported final values without baseline data. If a study directly provided the group mean and standard deviation of the change between baseline and the end of the study, we used those values. Otherwise, we calculated the group MD, and we derived the standard deviation (SD) by multiplying the standard error (SE) of the mean difference by the square root of the sample size (N), with SD = SE * √N. When separate data for two groups within the same study arm (e.g., low-risk and high-risk) were available but pooled data were required, we estimated the pooled mean and the pooled standard deviation or the pooled number of events, depending on the outcome type. The pooled mean was calculated as the weighted average of group means based on their sample sizes. The pooled SD was derived using the standard formula for combining standard deviations of independent groups, weighted by their respective sample sizes. SD pooled = √((N1-1) * SD1^2^ + (N2-1) * SD2^2^) / (N1 + N2-2), where SD1 and SD2 are the standard deviations of the two groups, and N1 and N2 are their respective sample sizes. When outcomes were reported as minimum, maximum, median, and first and third quartiles, we estimated the mean using the method proposed by Luo et al. (2018) (31) and the standard deviation using the method proposed by Shi et al. (2020) (32). Before applying these estimations, we verified whether the data were significantly skewed from normality using the formula described by Shi et al. (2023) (33). In cases where skewness was detected, we applied a logarithmic transformation to the summary data. After confirming that the transformed data were approximately normally distributed, we estimated the mean and standard deviation on the log-transformed scale and subsequently back-transformed the results to the original scale. For binary outcomes, the pooled number of events was calculated as the sum of events across both groups (E pooled = E1 + E2) and the pooled event rate was computed by dividing the total number of events by the total number of participants (E1 + E2) / (N1 + N2). Mean difference was calculated for all outcomes; those with data from only one study were reported descriptively, while outcomes with data from at least two studies were subjected to meta-analysis. We combined studies together using the inverse variance-weighted method and used both fixed and random-effect models (34). We used the restricted maximum-likelihood (REML) estimator to estimate the between-study variance (35). We tested for heterogeneity using the Cochran’s Q test and quantified it through the I^2^ statistic, interpreting values <50% as low, between 50% and 75% as moderate, and >75% (36) as high heterogeneity. We created forest plots for graphical data presentation, including the point estimate, 95% CI, and study descriptors (first author, year of publication). We performed all statistical data analyses using R version 4.4.0 (2024-04-24) for Windows and used the meta package for the meta-analyses.

## RESULTS

We retrieved 7,830 articles and after deduplication 5,971 articles were assessed. 204 articles were included after screening for title and abstract. 21 reports were protocols or registrations of unique RCTs. We contacted the 21 PIs of these included eligible protocols, however only 6 of them answered. Five of them responded that they were still in the enrolment phase and did not have any available data yet, and one responded that the article was submitted to a scientific journal and did not want to share the results before publication. The remaining 15 authors did not respond. 183 articles were assessed for full text review, and we included 33 reports (37–69) pertaining to 27 unique RCTs (39,44–69) in the final analysis. We excluded the 150 studies for the following reasons: 12 were secondary analysis of the included studies that did not provide any additional outcome; 52 did not follow a RCT design; in 22 studies PRS disclosure was not randomized among participants; 30 studies focused on genetic testing of single gene pathogenetic (non-common) variant rather than polygenic risk scores; 13 studies utilized information from only one common SNP; and 21 were protocols of the included studies (Fig. 1).

**Figure 1.**
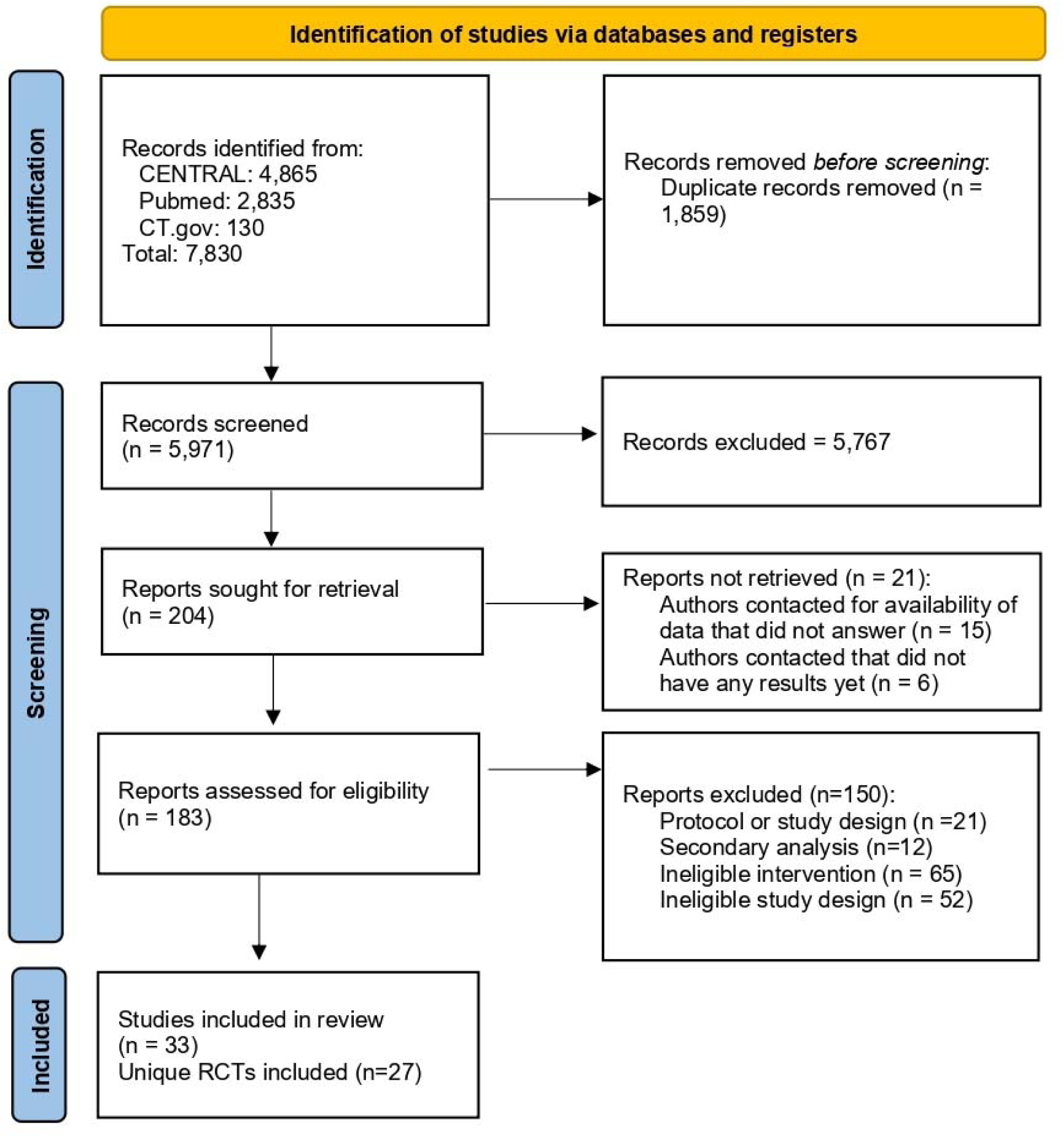
PRISMA flow diagram of the study search and selection proces.

### Otto-SR Validation

After reproducing the screening and extraction phases using the Otto-SR outputs, we found a high degree of comparability between the results obtained by human reviewers and those generated by the LLM.

For the screening phase, Otto-SR identified a total of 67 potentially eligible full-text articles. Four studies were not retrievable by Otto-SR, as they had been identified through had been identified through hand search of the main paper, starting from secondary publications included in the search string (39,40,56) and thorugh the ClinicalTrials.gov screening (51); therefore, the total number of eligible and retrievable reports for assessment was 29. Of the 29 eligible reports, otto-SR correctly included 27 reports (93.1% sensitivity); and humans correctly included 28 reports (96.5% sensitivity). Two articles (46,53) were incorrectly excluded by otto-SR due to the misinterpretation of the >2 SNP threshold for PRS classification. One article (58) was incorrectly excluded by human reviewers, but additionally captured by the otto-SR system.

Among the remaining 40 records included by otto-SR, 36 were secondary analyses without outcomes, potentially eligible congress abstracts missing key information for inclusion, protocols of included trials, or protocols for which we contacted the principal investigators (PIs). Thus, Otto-SR misclassified 6 articles (99.9% specificity) as eligible (false positive).

For the extraction of individual study data and general characteristics, a total of 270 fields were analyzed. Of these, 249 were correctly extracted by both the human reviewer and Otto-SR, corresponding to a concordance of 92.2%. Among the remaining discrepancies, 5 fields (1.9%) were extracted incorrectly by the human reviewer and 16 (5.9%) by Otto-SR.

For numerical outcome data extraction, 1,415 fields were examined. Otto-SR achieved a concordance of 98.6%, with 16 fields (1.1%) extracted incorrectly compared with 4 (0.3%) by the human reviewer.

### Characteristics of the studies

Table 1 presents general information about the 27 included RCTs. The median sample size across studies was 207, with a range of 67 to 3,177 participants. Most studies were conducted after 2017 (n = 17 studies) and showed variable geographical distribution, with the United States hosting half of studies (n = 11), followed by Europe (n=5) and United Kingdom (n = 3). The follow-up times of participants across studies had a median of 6 months, ranging from 1 to 12 months. Twenty four studies were registered on a registry, with 17 on ClinicalTrial.gov and 7 on other registries (e.g. ISRCTN or ANZCTR).

**Table 1.**
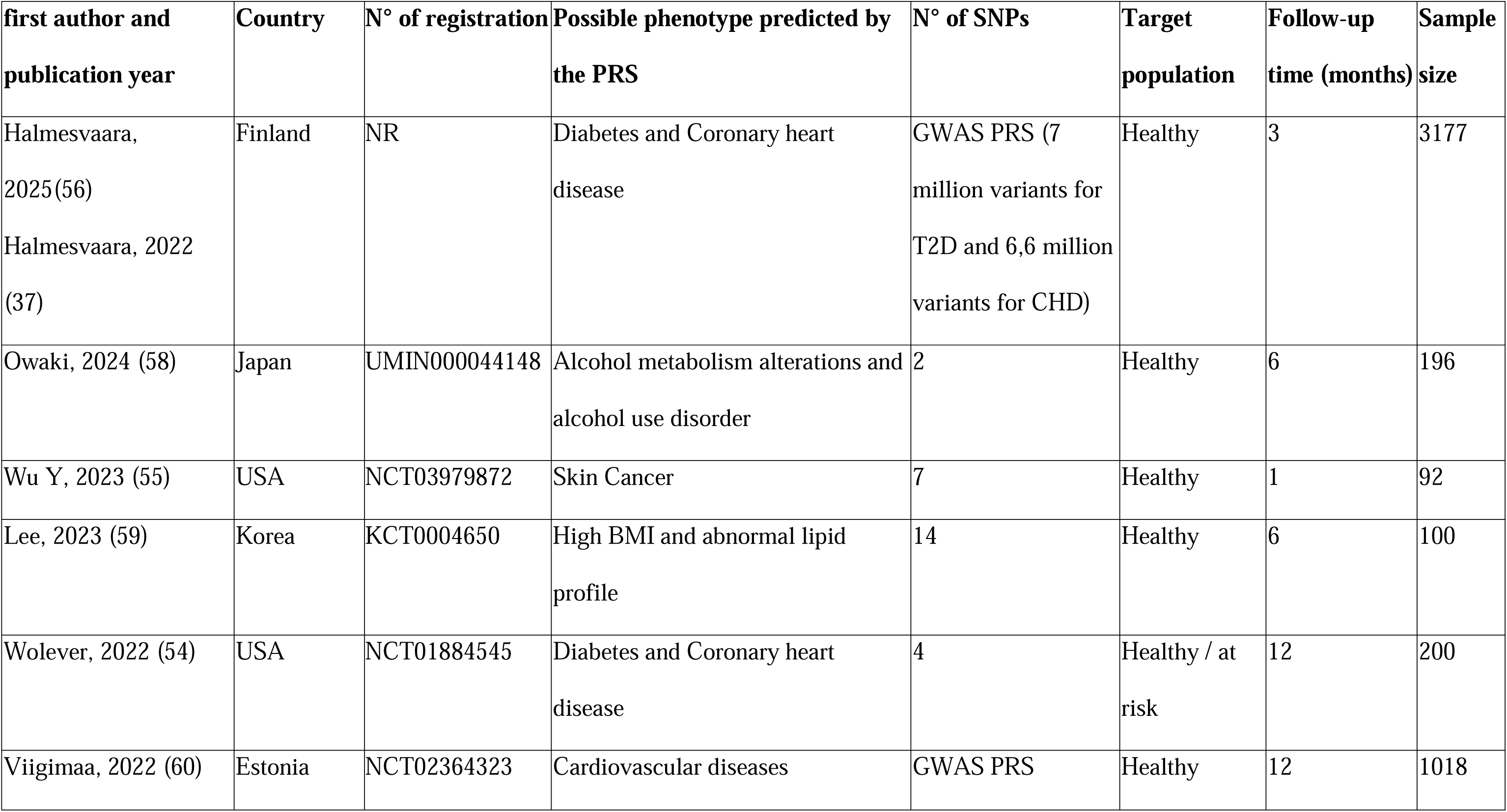
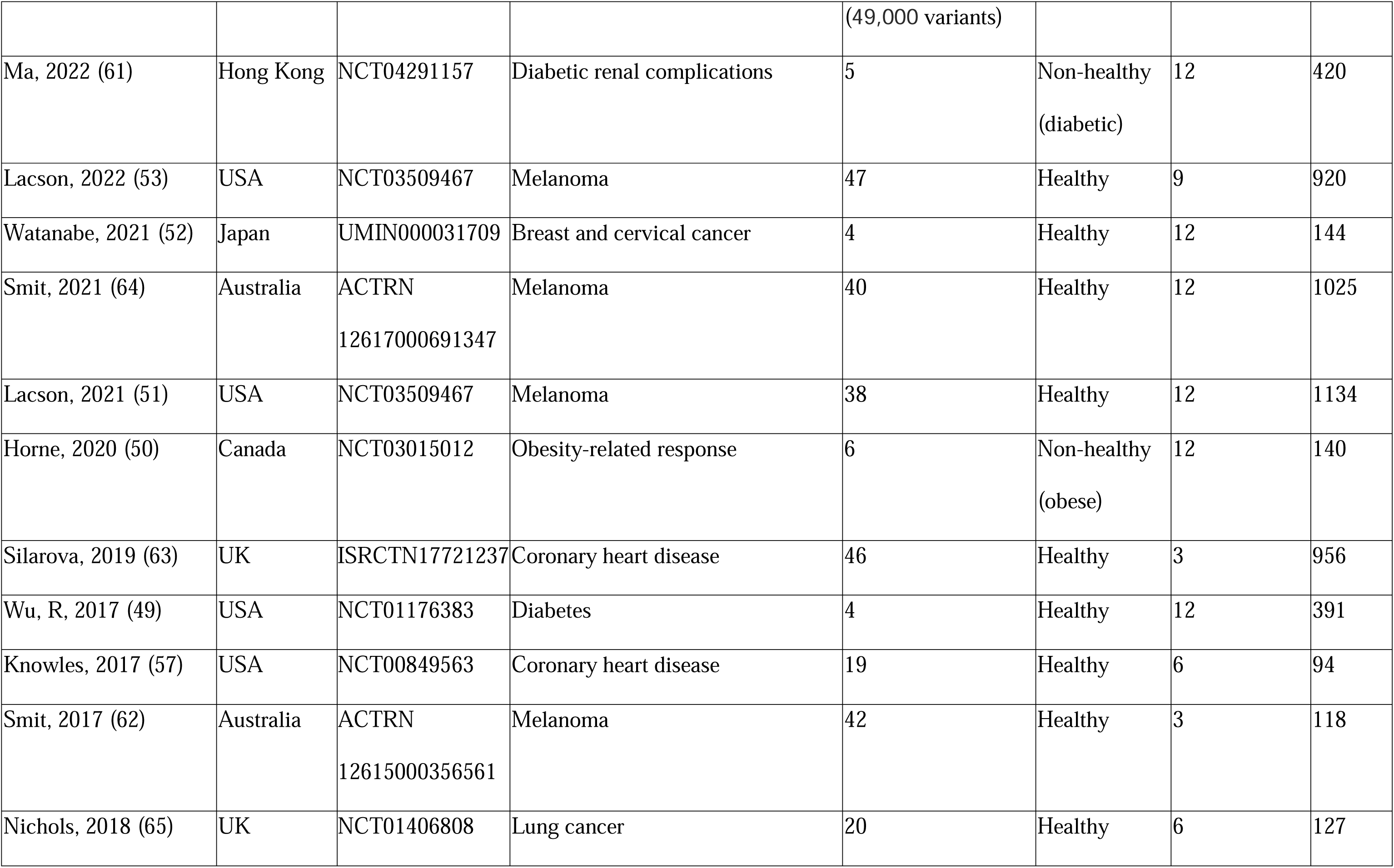
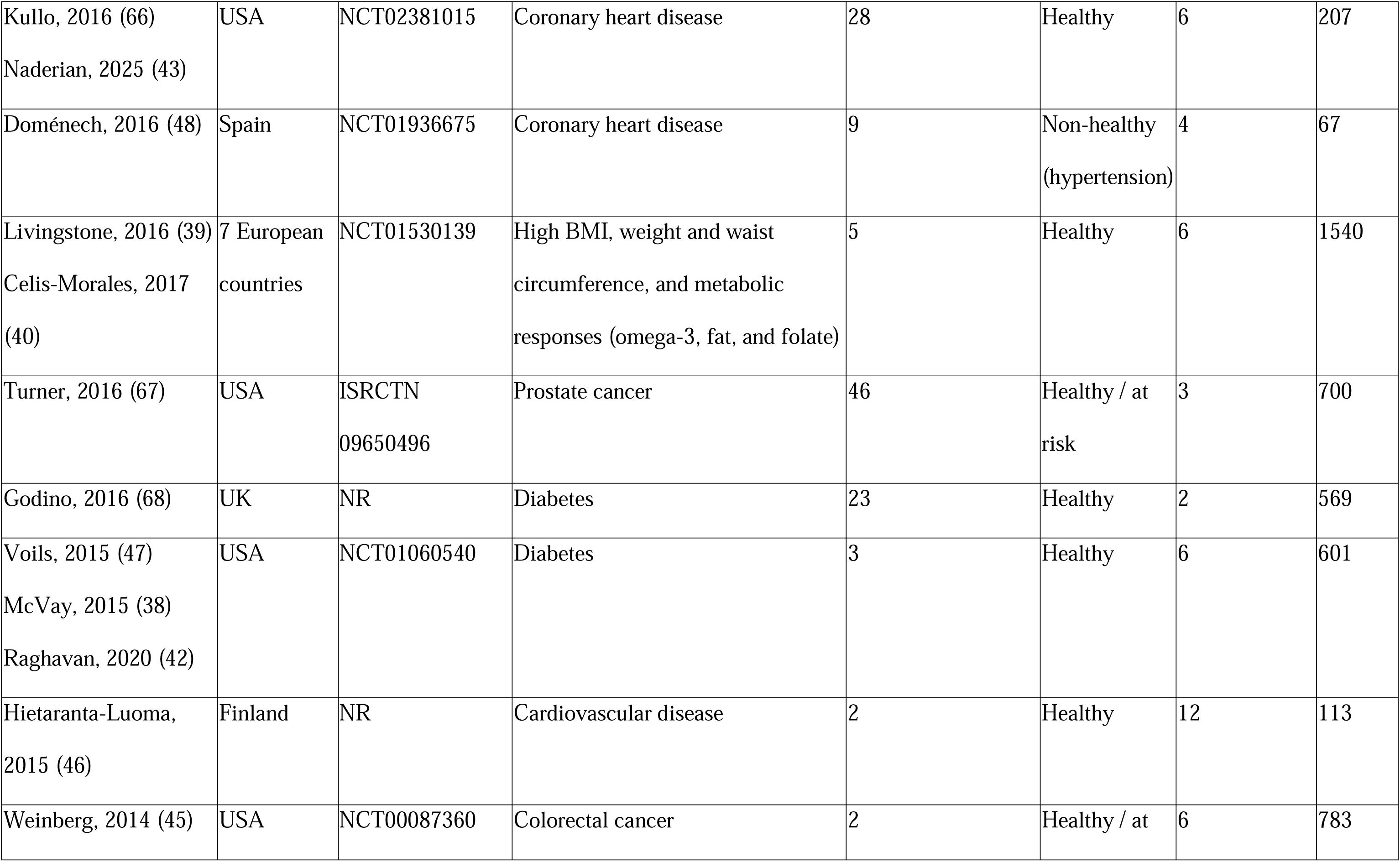
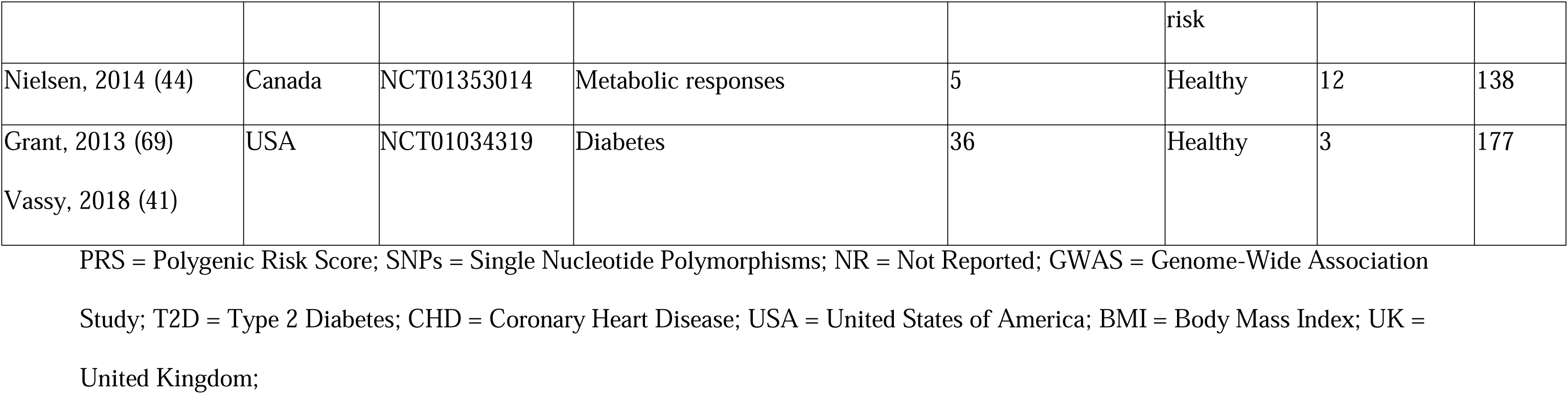
Characteristics of the 27 RCTs included in the systematic review.

Regarding the phenotypes predicted by the PRS, cancers (n=9) and cardiovascular diseases (CVDs) (n=8) were the most common, followed by diabetes (n = 6). In two studies, the PRS predicted both diabetes and CVDs. Other predicted phenotypes included metabolic alteration (n=4) including high BMI and abnormal lipid profile, metabolic responses, obesity-related response, and alcohol metabolism, while one study predicted the development of diabetic renal complications. Among cancer-related studies, melanoma (n = 5), was the most frequently investigated.

Twenty one studies focused on healthy (n=21) or at-risk (n=3) populations, while three studies targeted individuals who had already developed a disease. In these three studies (Ma et al., Horne, 2020, Doménech, 2016), the PRS was used to predict disease complications. Specifically, in Ma et al., the PRS predicts the risk of developing diabetes-related complications, particularly kidney disease; in Doménech et al., it predicts the risk of cardiovascular disease in patients already diagnosed with hypertension; and in Horne et al., it predicts the response to weight loss and physical activity in obese individuals.

Table 2 reports the details of the interventions and outcomes of the RCTs. Twenty one studies were two-arm RCTs (77.8%), while the remaining studies were three (n=1) or four (n=5) arm trials, including comparisons with other estimated phenotypic risk or other information, like web-based advice. The studies evaluated interventions involving the communication of PRS, either alone (n = 3) or combined with additional or combined strategies, including personalized booklets or educational materials (n = 7), genetic counseling (n = 5), lifestyle advice (n = 2), or behavioral programs such as smoking cessation or diabetes prevention (n = 2). Specifically, the behavioral programmes were given also to the control arm. The information in the control arms was usually traditional risk assessment (n = 7), standard of care (n = 12), or no information or intervention (n = 4). In six of the 2-arm RCTs, the intervention group was further subclassified into an intervention-low-risk group and an intervention-high-risk group.

**Table 2.**
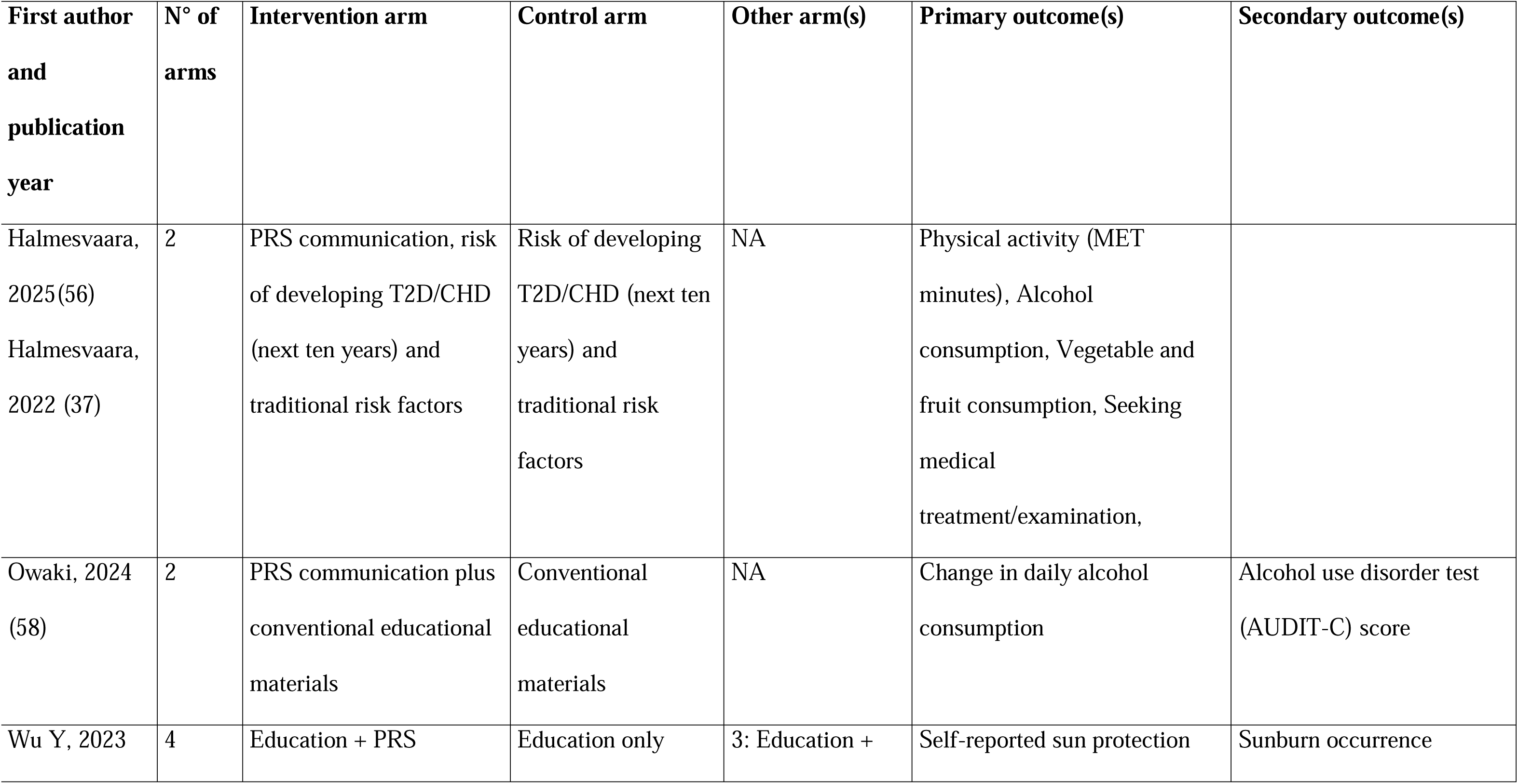
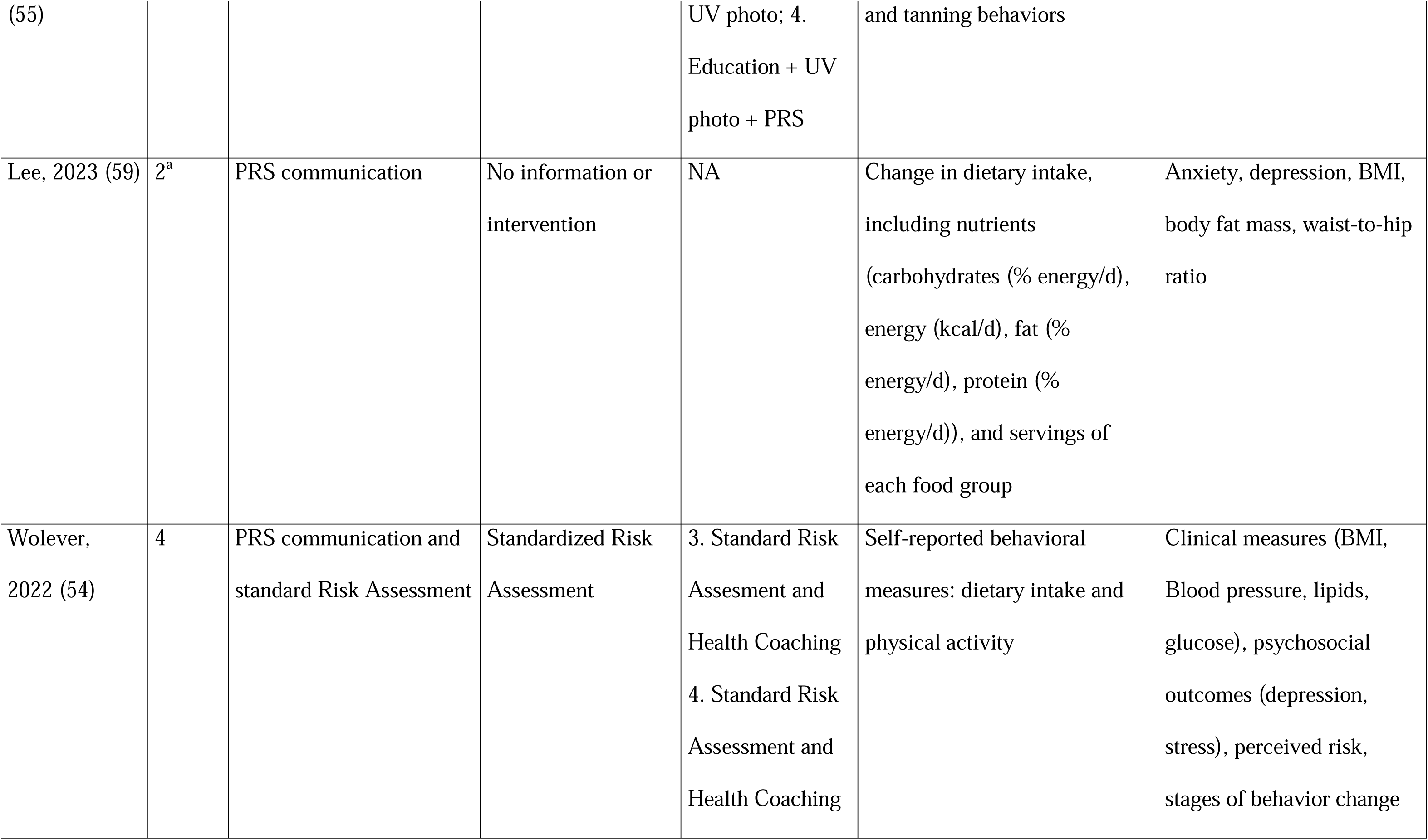
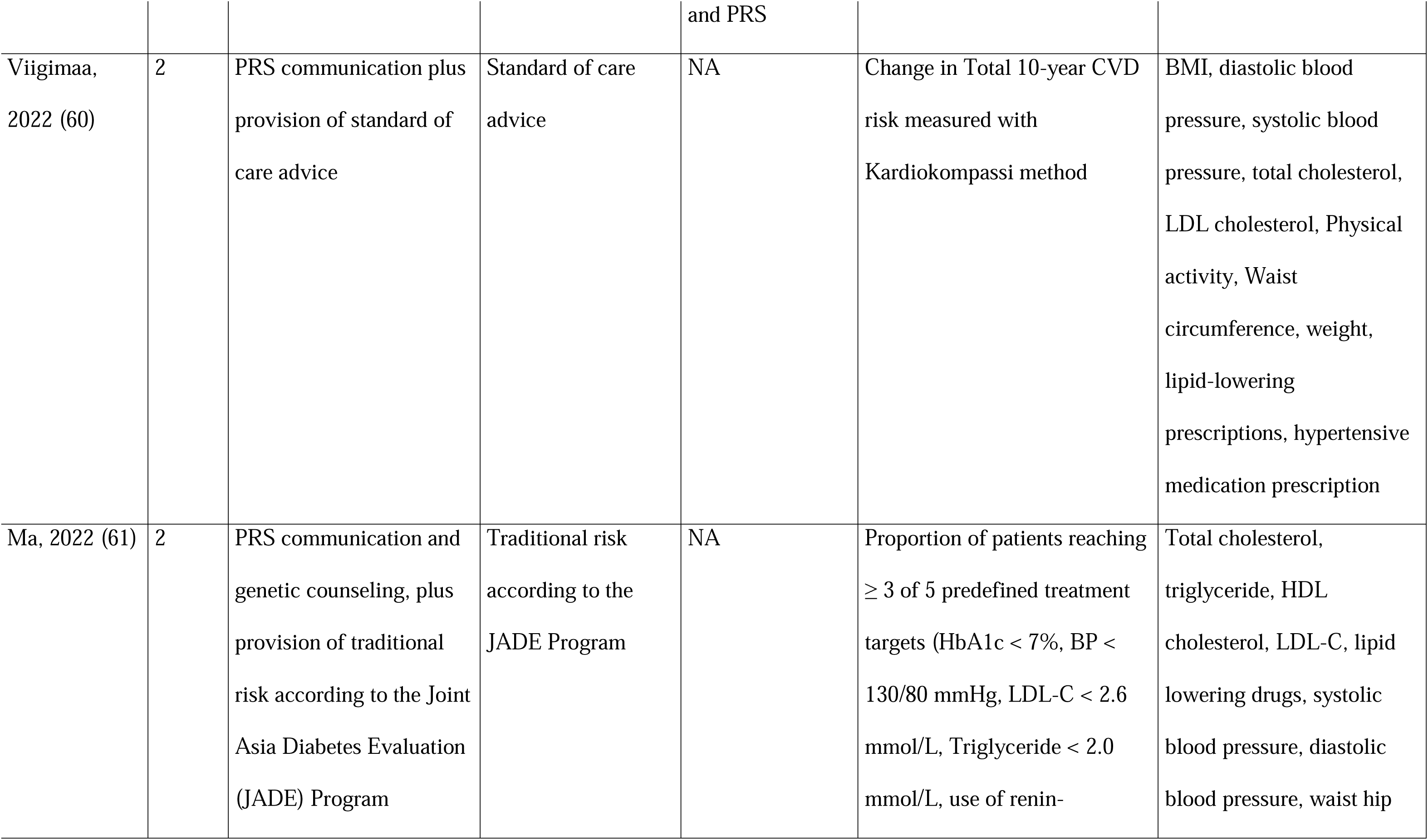
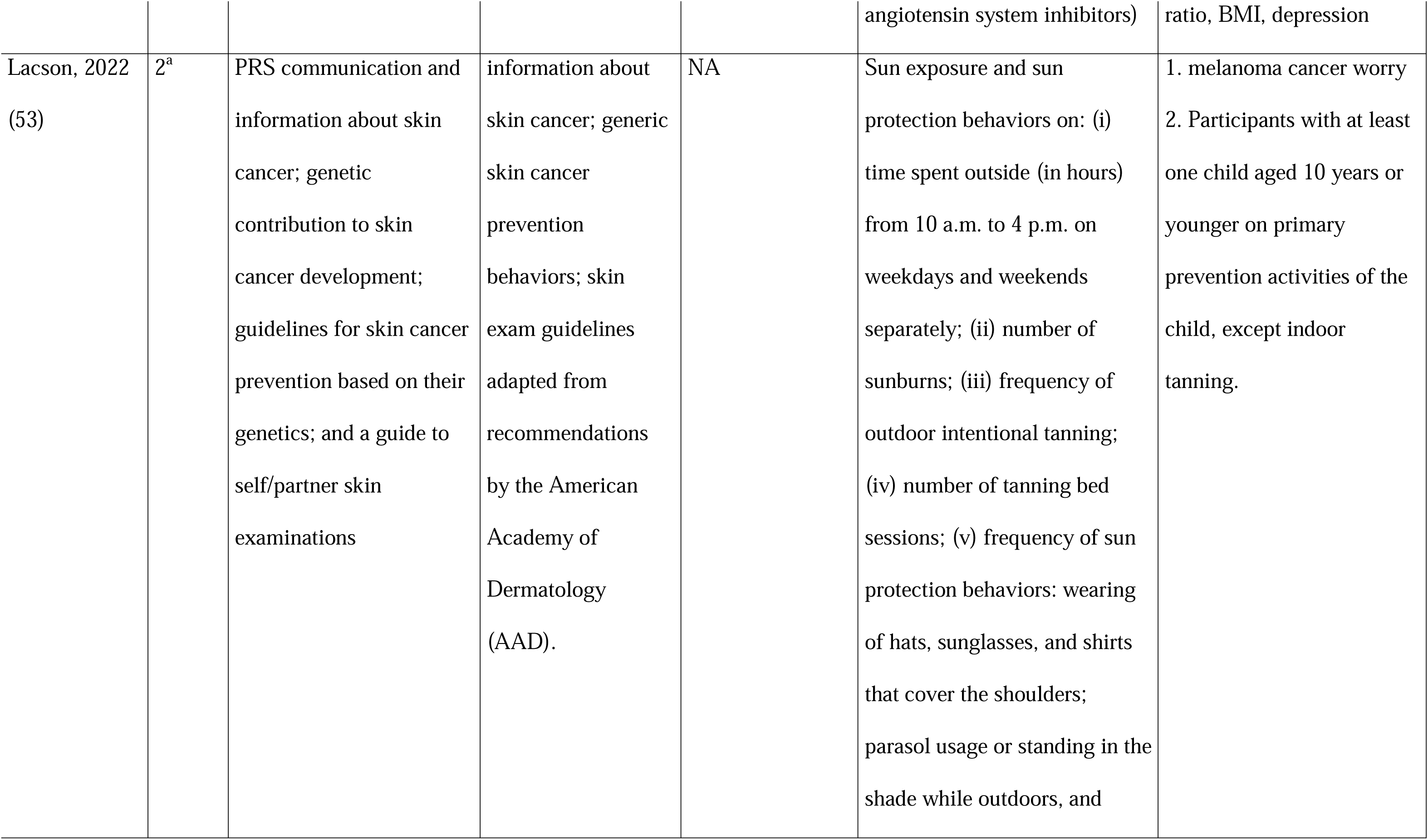
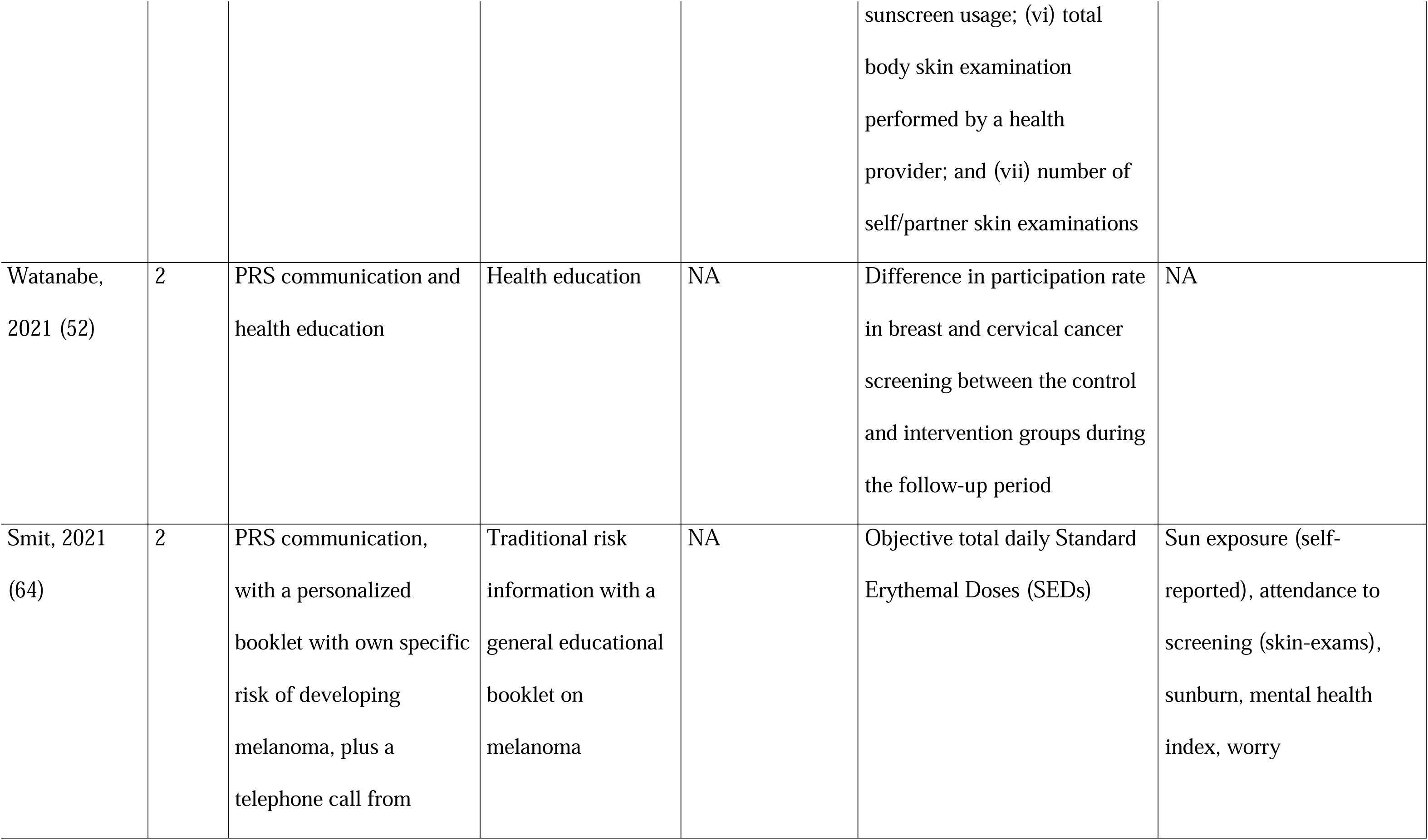
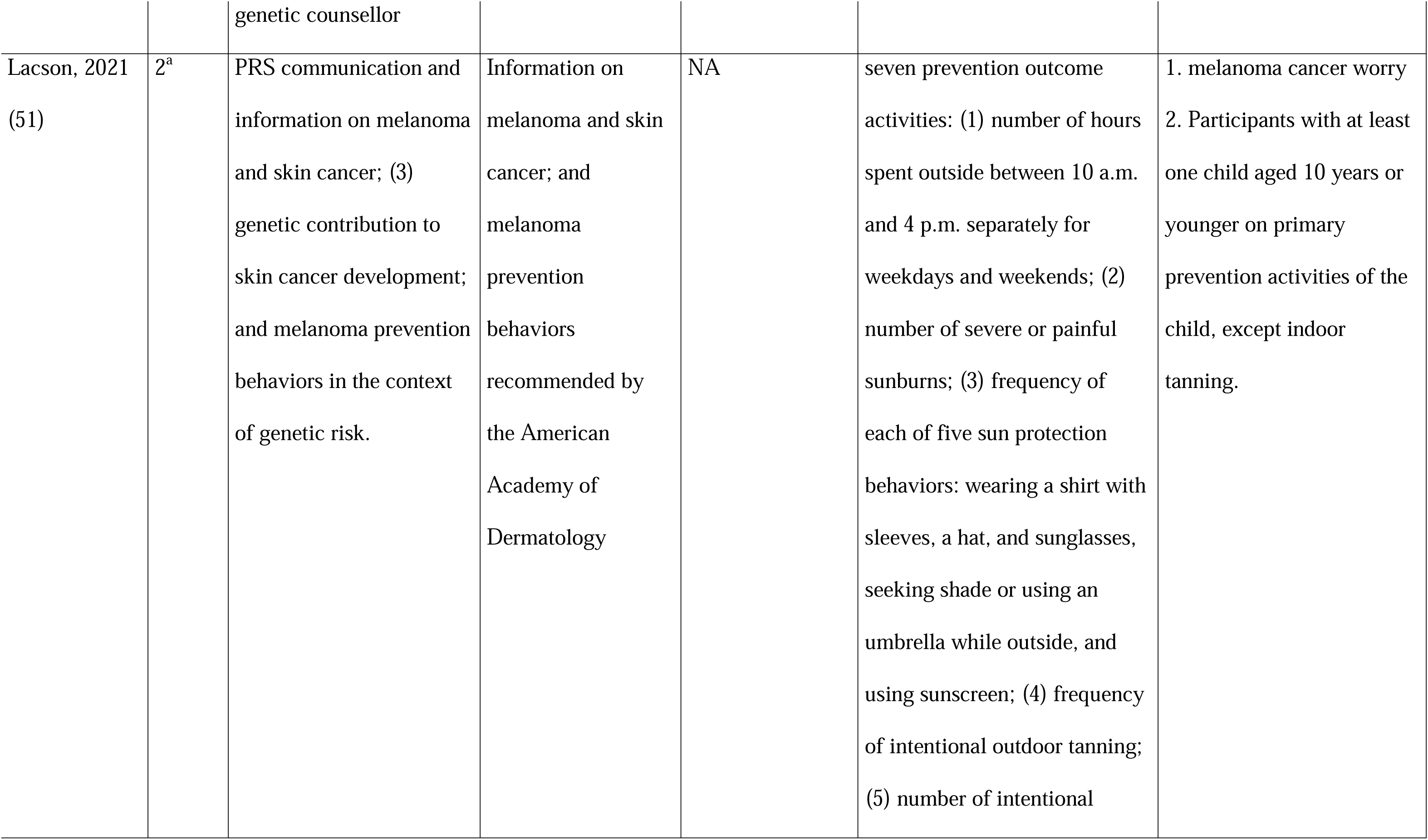
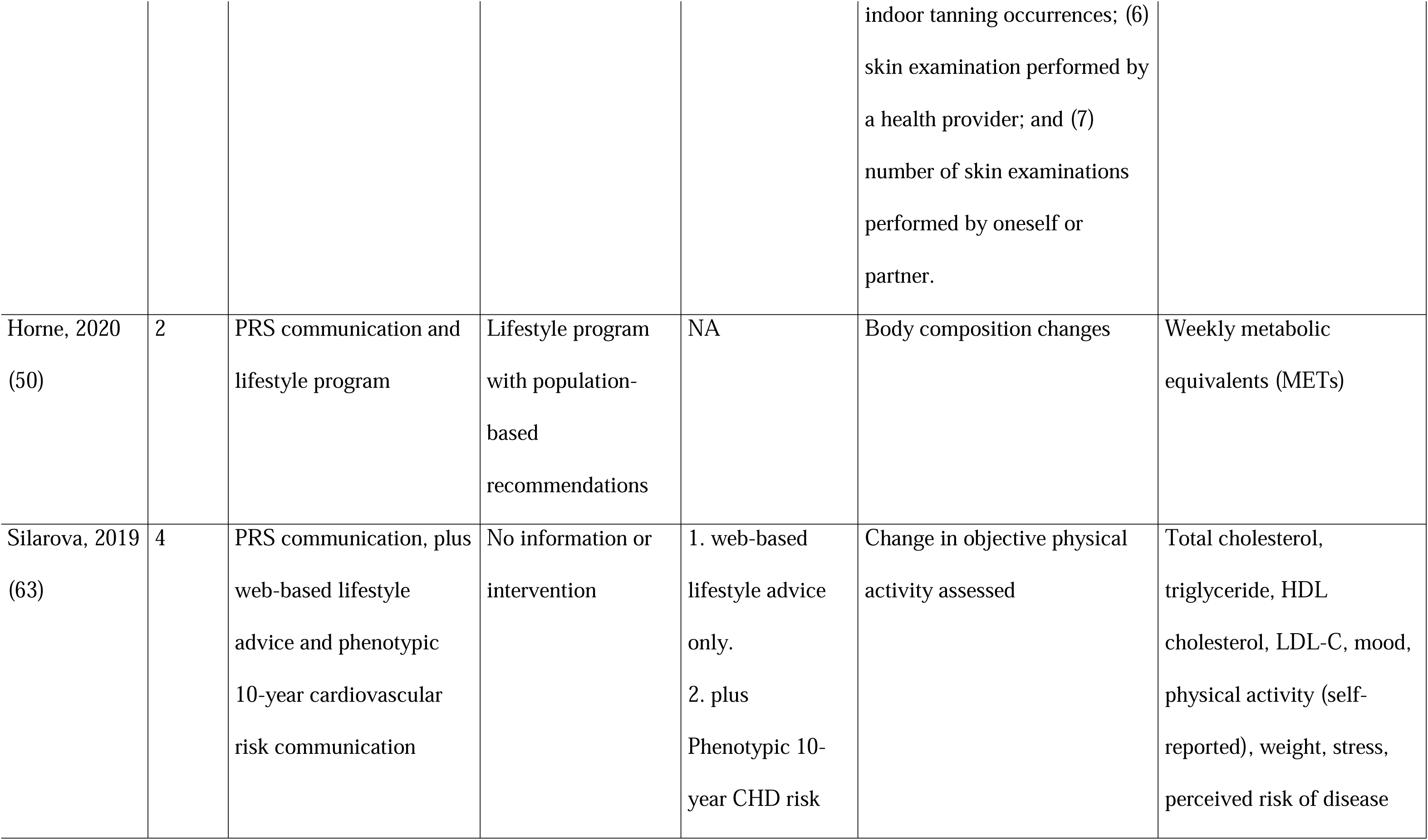
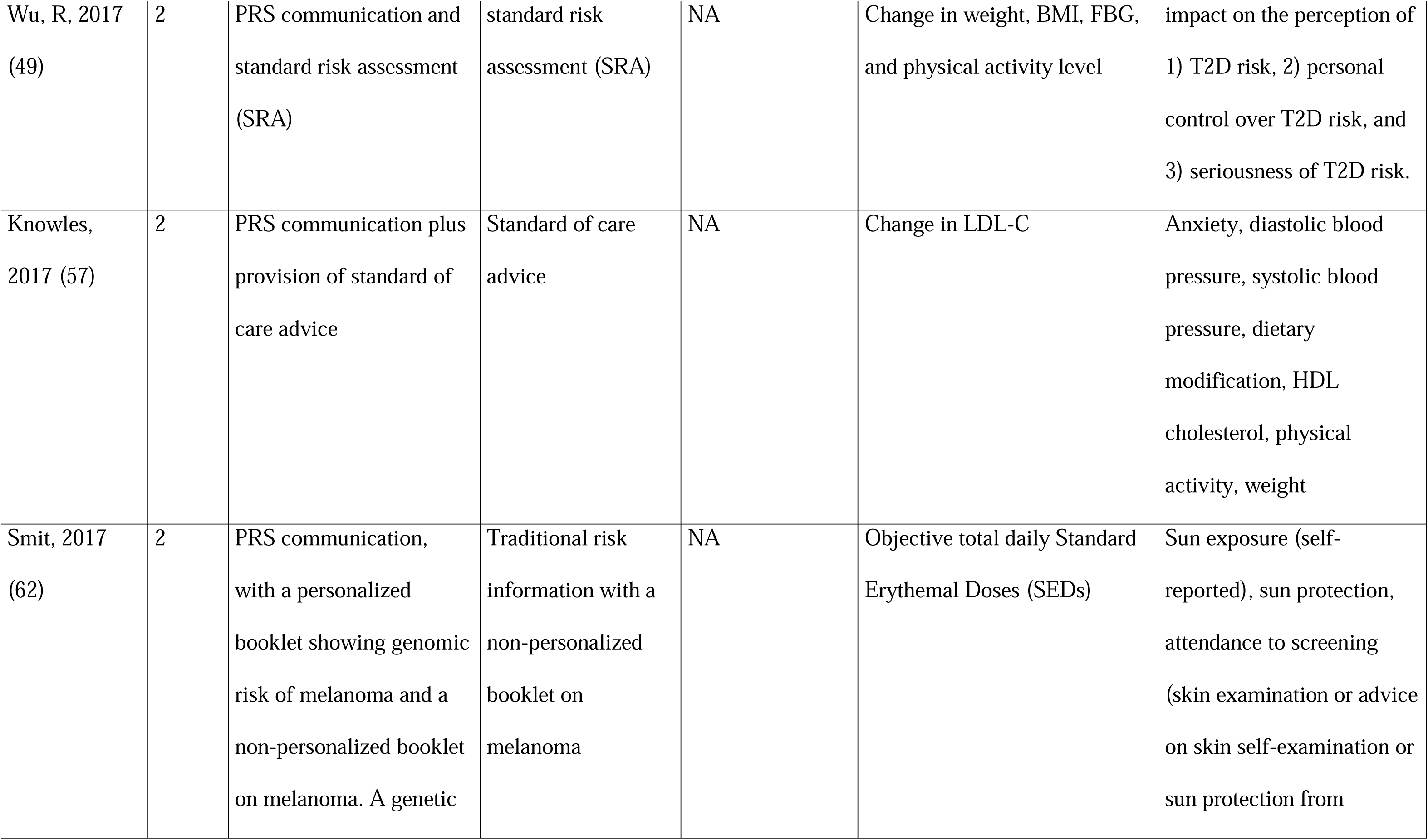
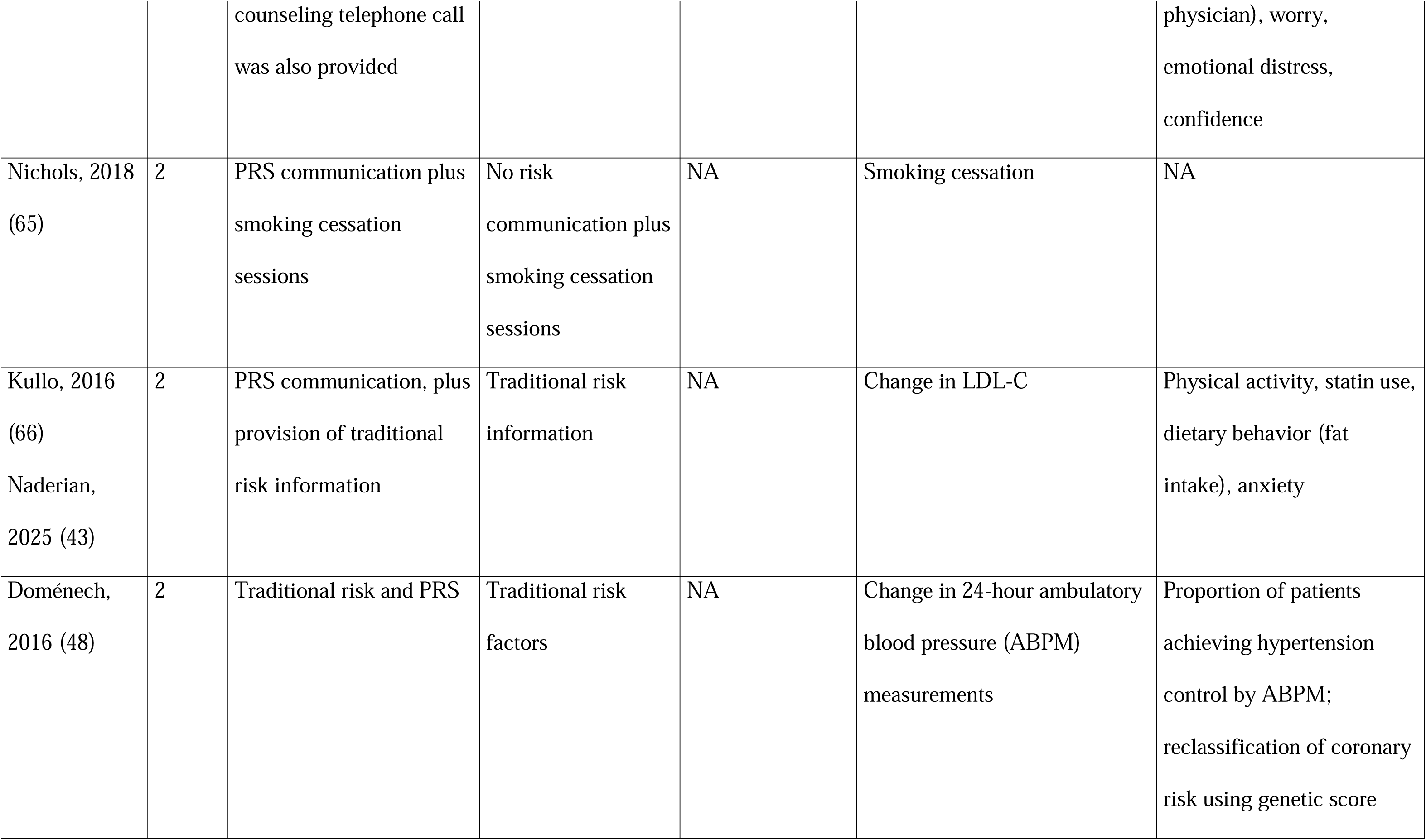
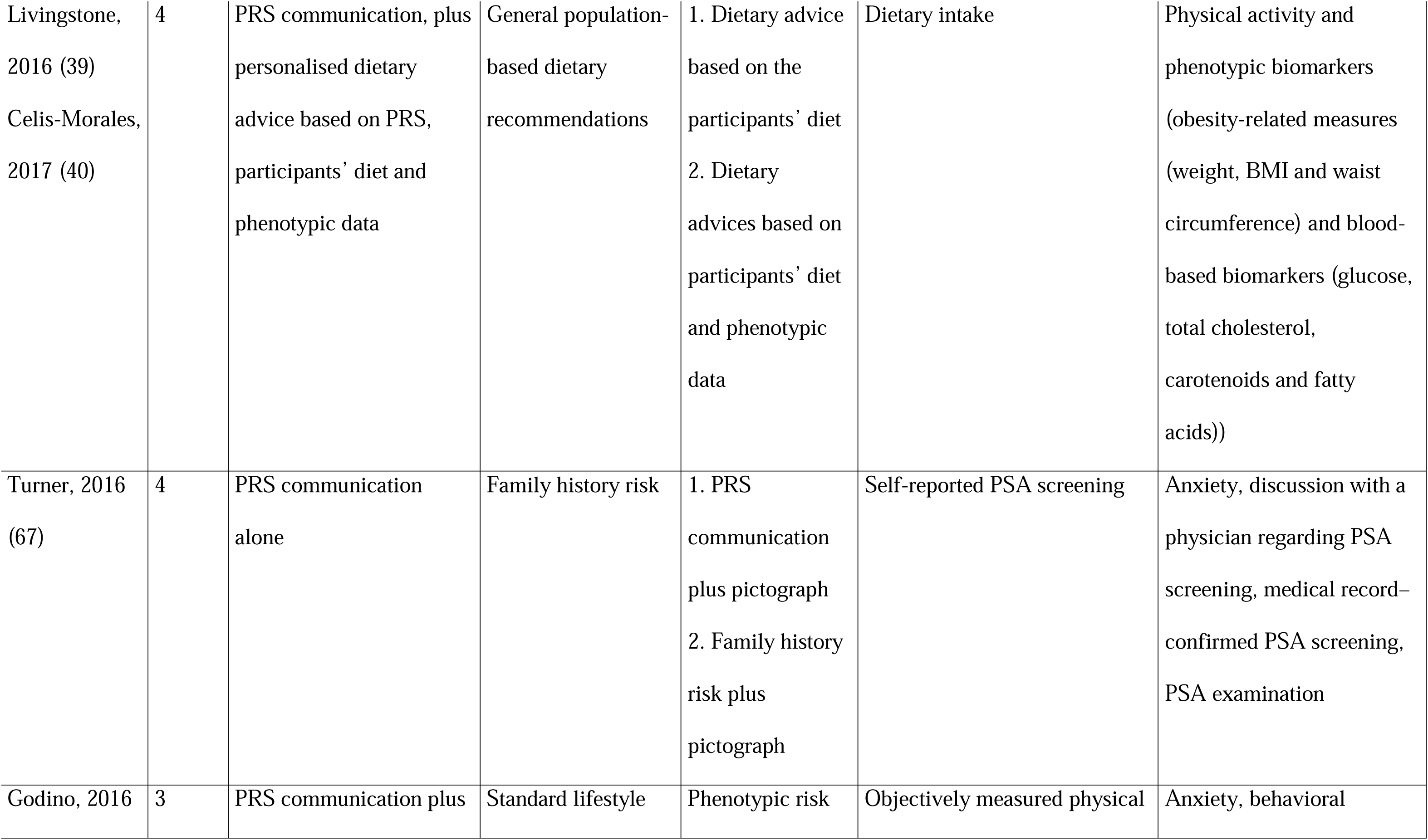
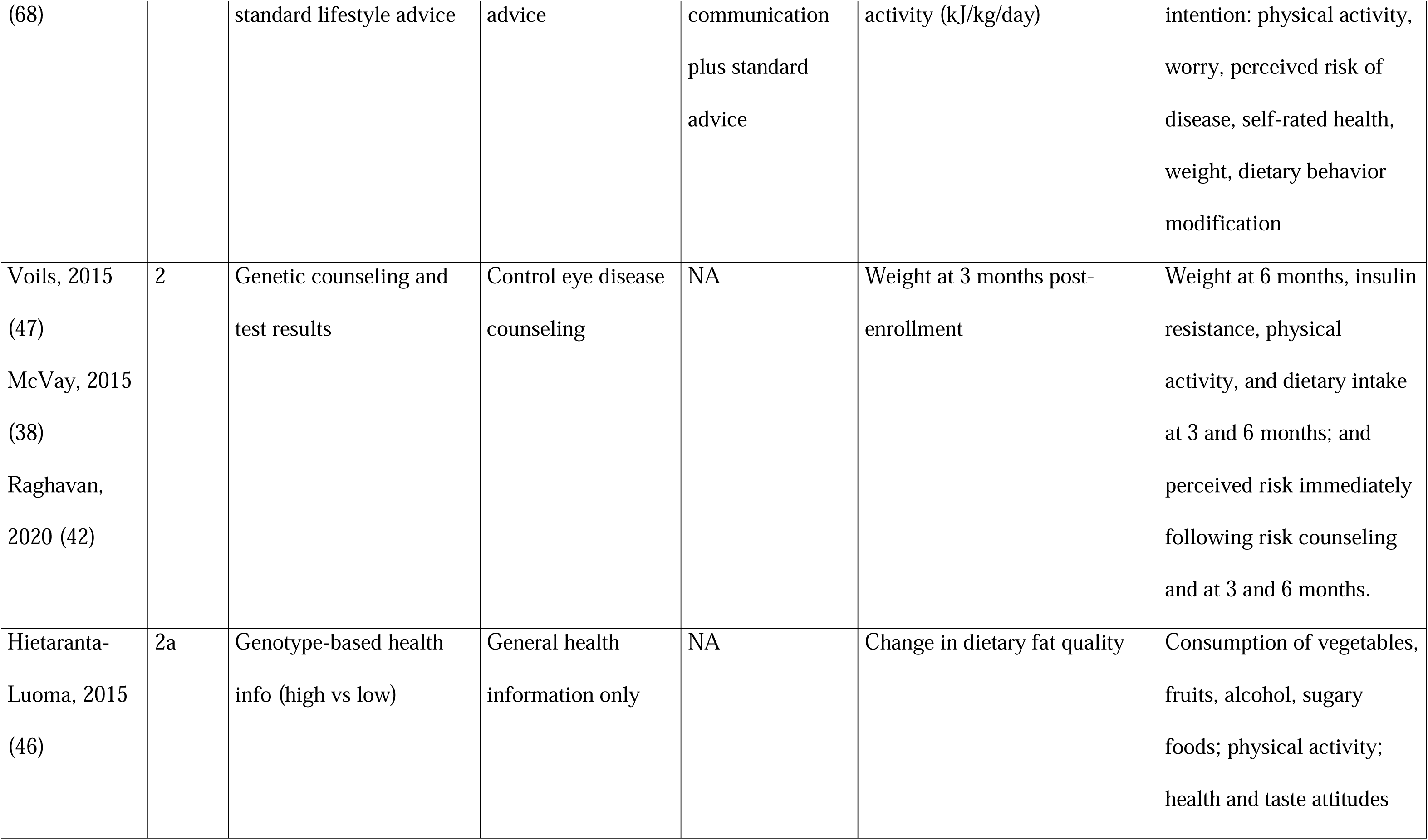
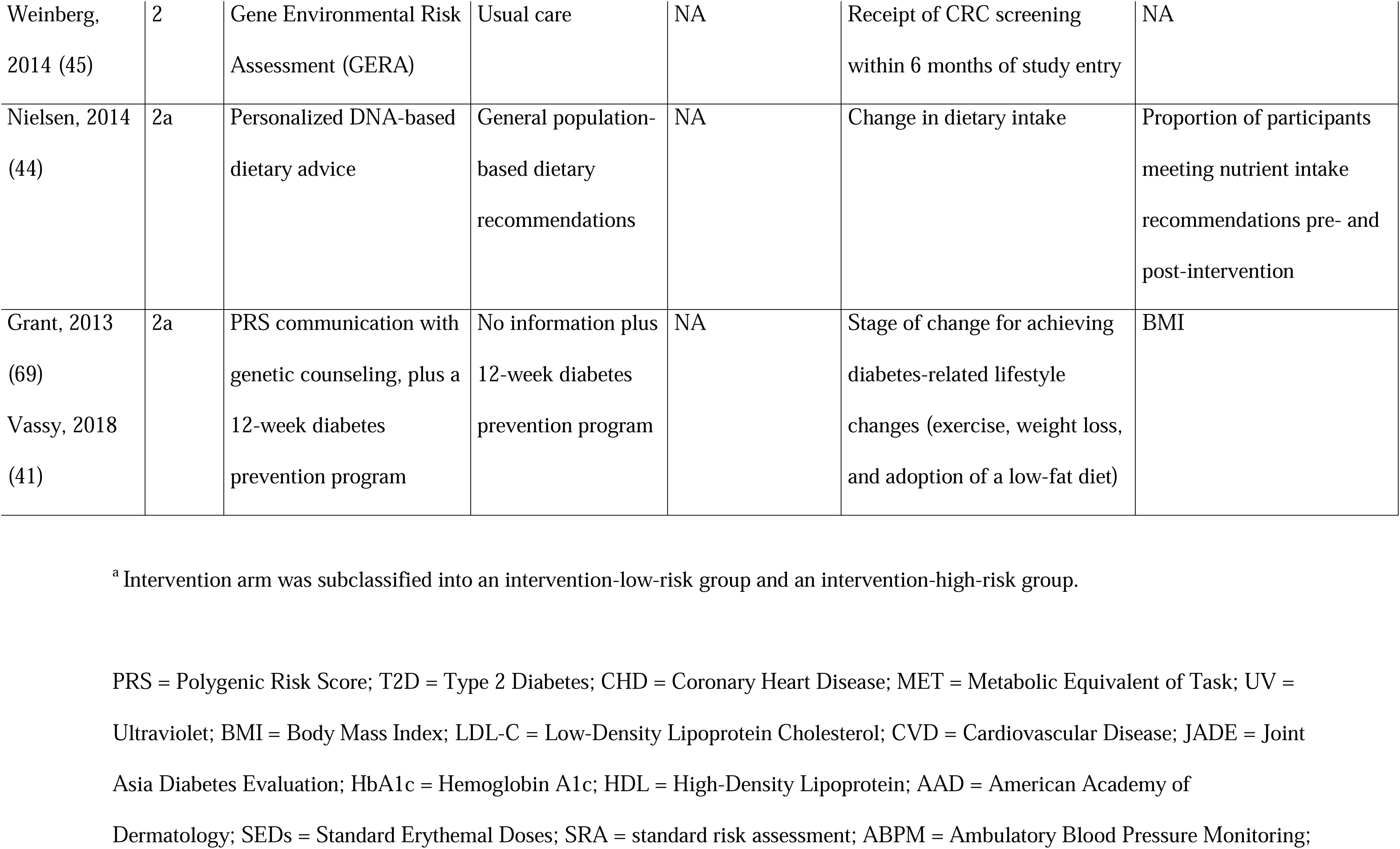
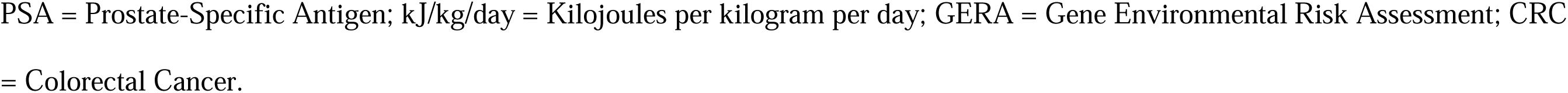
Intervention and outcome characteristics of the 27 RCTs included in the systematic review.

### Risk of bias (RoB-2) evaluation

The results of the risk of bias evaluation are reported in Fig. 2. Overall, 9 studies (33.3%) were assessed at high risk for overall bias, 11 (40.8%) raised some concerns, and 7 (25.9%) were at low risk. Missing outcome data was the most frequent domain contributing to high risk of bias.

**Figure 2.**
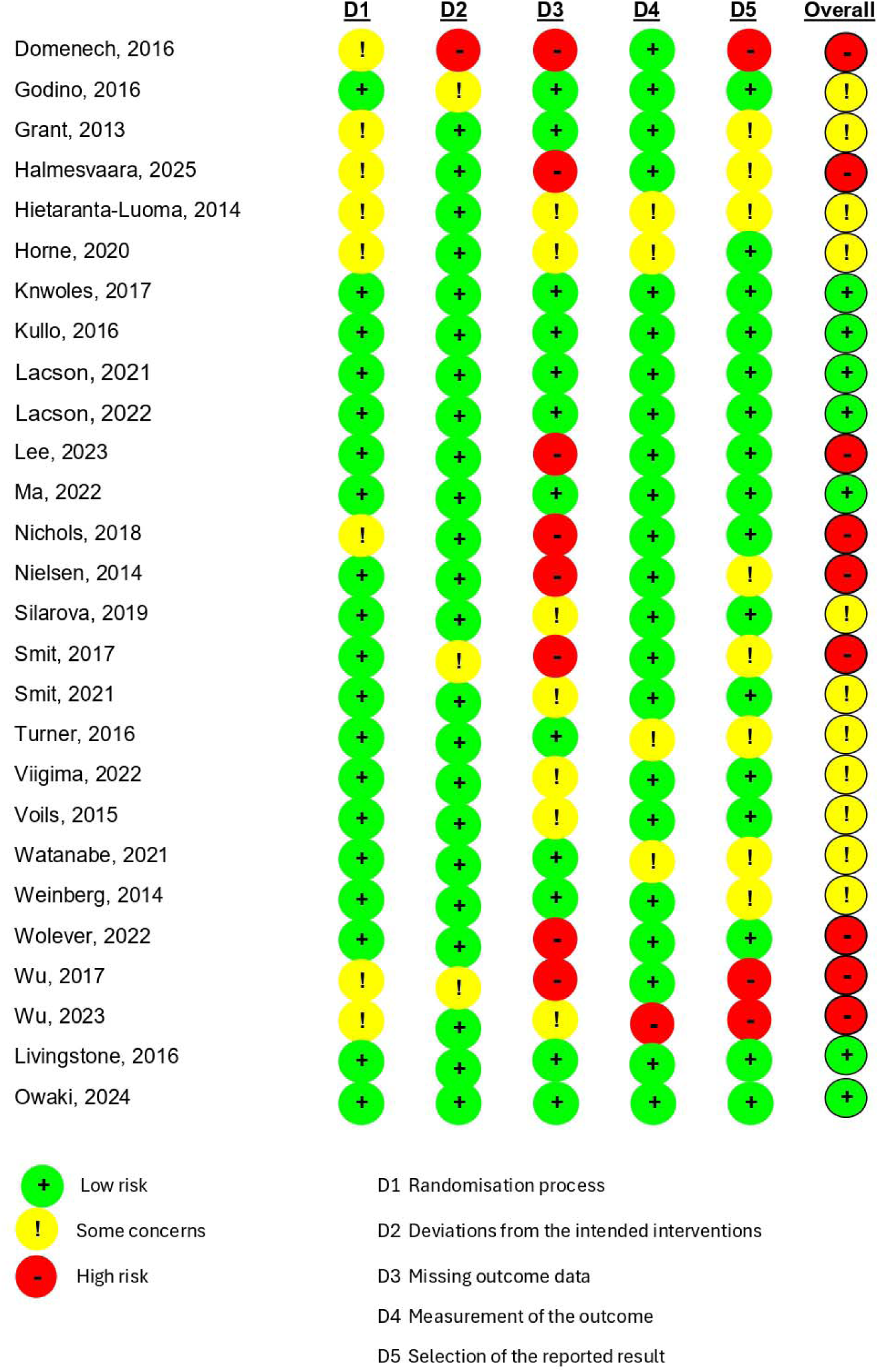
Risk of bias with RoB-2 tool of the 27 trials included.

### Outcomes with meta-analysis

Table 3 reports the results of the meta-analysis for 22 outcomes, while forest plots are available in Supplementary file 1. The meta-analyses revealed no statistically significant effects across the assessed outcomes. These outcomes included lipids, weight, blood pressure, physical activity, dietary and sun related behaviors, and clinical and psychological outcome. Moreover, the point estimates were close to the null, with SMD estimates ranging from −0.11 to 0.08, RR estimates falling <1.5, and relatively small mean differences in lipid, weight, and blood pressure outcomes.

**Table 3.** Meta-analysis of differences from baseline to final follow-up in the 27 RCTs included.

| Outcome | Measure | N° of studies (intervention; control) | Value | 95%CI | I<sup>2</sup> | References |
| --- | --- | --- | --- | --- | --- | --- |
| LDL cholesterol | Mean difference | 4 (514; 537) | -3.64 | [-7.88; 0.60] | 0% | (57,61,63,66) |
| HDL cholesterol* | Mean difference | 3 (411; 437) | -0.21 | [-2.65; 2.23] | 1.7% | (57,61,63) |
| Total cholesterol* | Mean difference | 2 (391; 391) | -2.01 | [-8.27; 4.26] | 0% | (61,63) |
| Diastolic Blood Pressure | Mean difference | 3 (247; 276) | -1.88 | [-4.17; 0.42] | 0% | (48,57,61) |
| Systolic Blood Pressure | Mean difference | 3 (247; 276) | -1.26 | [-4.44; 1.92] | 10.1% | (48,57,61) |
| Weight | Mean difference | 4 (482; 443) | -0.33 | [-0.87; 0.20] | 0% | (57,63,68,69) |
| BMI | Mean difference | 3 (319; 271) | -0.12 | [-0.64; 0.39] | 0% | (59,61,69) |
| Physical activity | SMD | 4 (508; 511) | -0.01 | [-0.13; 0.11] | 0% | (57,63,66,68) |
| Diet (energy per day) | SMD | 3 (569; 531) | -0.11 | [-0.23; 0.01] | 0% | (59,68) |
| Diet (fat per day) | SMD | 2 (168; 133) | 0.08 | [-0.15; 0.31] | 0% | (59,66) |
| Alcohol consumption | SMD | 2 (277; 266) | -0.11 | [-0.28; 0.06] | 0% | (58,63) |
| Intentional tanning | SMD | 4 (1109; 1120) | -0.03 | [-0.11; 0.06] | 0% | (51,55,62,64) |
| Peak sun exposure | SMD | 2 (533; 548) | 0.02 | [-0.10; 0.14] | 0% | (62,64) |
| Sun Protection | SMD | 2 (533; 548) | 0.04 | [-0.08; 0.16] | 0% | (62,64) |
| Total Sun exposure | SMD | 3 (1091; 1101) | 0.00 | [-0.09; 0.08] | 0% | (51,62,64) |
| Incidence of the disease | Relative risk | 3 (467; 418) | 0.95 | [0.32; 2.79] | 68.3% | (41–43) |
| Screening attendance | Relative risk | 3 (990; 748) | 1.12 | [0.77; 1.61] | 25.5% | (45,52,67) |
| Statin usage* | Relative risk | 3 (760; 791) | 1.50 | [0.98; 2.29] | 88.7% | (60,61,66) |
| Depression* | SMD | 2 (245;237) | -0.05 | [-0.23; 0.13] | 0% | (59,61) |
| Perceived risk* | SMD | 2 (379; 379) | -0.04 | [-0.21; 0.13] | 31.1% | (63,68) |
| Anxiety* | SMD | 5 (702; 671) | -0.02 | [-0.13; 0.08] | 0% | (57,59,66–68) |
| Worry* | SMD | 3 (718; 733) | -0.06 | [-0.23; 0.10] | 42.4% | (62,64,68) |

\*=in all studies, those were secondary outcomes.
LDL = Low-Density Lipoprotein; HDL = High-Density Lipoprotein; BMI = Body Mass Index; SMD = Standardized Mean Difference.

Meta-analyses of the final value and the sensitivity analyses did not reveal any major modifications, as shown in supplementary tables 1, 2 and 3. Meta-analyzed forest plots of final value are available in Supplementary file 2.

### Narrative description of outcomes reported in single studies

Supplementary table 4 reports the analysis for 69 outcomes outcomes across multiple studies, with each outcome reported only once. Of these, 18 had been presented as primary outcomes in the respective RCTs, and the others as secondary.

Among the 18 primary outcomes, two had a statistically significant change. Ma et al. (61) reported a moderate improvement for its single primary outcome, change in composite treatment target score for diabetes (RR = 0.84; 95% CI: 0.70; 0.99). Such score was a composite end-point defined as the proportions of patients attaining ≥ 3 of 5 pre-defined treatment targets: 1) BP < 130/80 mmHg; 2) HbA1c < 7%; 3) LDL-cholesterol < 2.6 mmol/l; 4) Fasting triglyceride < 2.0 mmol/l and 5) Use of RASi for renoprotection. Livingstone et al. (39), found a small improvement for its single primary outcome, change in the MedDiet score (mean difference = +0.24; 95% CI: 0.08, 0.39), defined as adherence to the Mediterranean diet. Among the 51 secondary outcomes analyzed, only 2 had statistically significant differences (55,62). There was an increased confidence, measured with the question “How confident are you in your ability to identify melanoma? (from 1 “not at all” to 5 “very”)” in identifying melanoma in Smit 2017 et al. (62) (MD = 0.40; 95% CI: 0.10; 0.69); and also a decrease in reported sunburn occurrence in the past month assessed using an item from the Sun Habits Survey (from “0 times” to “5 or more times”) over the past month in Wu Y et al. (55) (MD = –0.81; 95% CI: –1.57; –0.05). No statistically significant results were reported for any other outcomes.

### Consistency between articles conclusions and reported results

Among the 27 RCTs, 15 concluded in favor of the genetic intervention, 5 presented mixed conclusions and called for further research, and 7 concluded not in favor of the intervention, as reported in supplementary table 5. However, among the 15 studies that concluded in favor, only five reported results that were both favorable and statistically significant. Specifically, Kullo et al. (66) reported a significant reduction in LDL-C values in the intervention group compared to baseline; Viigimaa et al. (60) observed significant changes in most of the analyzed outcomes (i.e. LDL and total cholesterol, hypertensive medication) in the intervention versus control groups; Horne et al. (50) reported a significant increase in physical activity; and Domenech et al. (48) reported significant changes in blood pressure. Livingstone et al. (39), also reported an improvement in dietary score.

In contrast, the remaining 10 RCTs that concluded in favor did not show any statistically significant effects in their main outcomes. In these cases, effects were limited to specific subgroup analyses or to short-term improvements. Three studies reported effects only among participants classified as high genetic risk; two study found differences exclusively within-group; two studies observed effects both in high-risk individuals and within-group comparisons; two studies reported effects restricted to other subgroups (e.g., males); and one study found a significant effect at ten weeks that was not sustained at follow-up.

## DISCUSSION

This systematic review and meta-analysis of 27 RCTs provides the most comprehensive synthesis to date of the effects of PRS communication on health-related outcomes. Across 22 outcomes meta-analyzed, no significant effects emerged for behavioral changes, including diet, physical activity, smoking, or screening attendance; psychological measures, including anxiety, depression, or risk perception; or clinical endpoints, namely lipid levels, blood pressure, BMI, or weight. A small number of individual trials reported favorable results, often restricted to short-term or subgroup analyses, but these findings were not consistent across studies and did not translate into significant summary effects when combined with other studies. Overall, despite frequent claims of benefit, the evidence does not support the conclusion that PRS disclosure meaningfully changes patient behavior, psychological well-being, or clinical outcomes.

This review used a standard protocol and followed standard methods and reporting guidelines. This approach enabled meta-analysis across a wide breadth of outcomes. Additionally, the artificial-intelligence platform Otto-SR was used to validate both the retrieval of eligible studies and data extraction values, to ensure that no potentially relevant trials or information were overlooked, resulting in very high concordance for data extraction and leading to the retrieval of an additional trial, missed during the screening phase by the human reviewers.

Nevertheless, several limitations should be acknowledged. Considerable heterogeneity was present in the way PRSs were constructed and communicated. Only a minority of studies used modern genome-wide PRSs including millions of variants, while most relied on a limited number of SNPs. Nearly one-third of trials were judged to be at high risk of bias, often due to missing outcome data. Sample sizes were generally modest, and follow-up periods were short (median six months), limiting the ability to detect any long-term effects and posing limits to the assessment of clinical outcomes that matter. Outcomes were frequently self-reported, particularly for diet and physical activity, which reduces reliability. Moreover, heterogeneous questionnaires and scales were used across studies, further limiting comparability. In several trials, baseline and final values were not consistently reported, with some studies presenting only differences or final measurements, thereby restricting the number of studies that could be meaningfully compared.

Our findings align with previous systematic reviews of genetic risk communication, which also reported minimal impact on health behaviors or psychological outcomes (19–21). However, earlier reviews frequently included single-gene variants or even pathogenic mutations, whereas this review focused specifically on PRSs and has many more RCTs included that examined PRSs rather than single genetic variants. Despite the additional genetic information, the evidence was generally not supportive of substantive benefits. This review emphasizes the persistent gap between the theoretical promise of PRS-guided prevention and its currently limited real-world effectiveness.

Several factors may explain the lack of meaningful benefit from PRS disclosure. First, patients may struggle to understand or act upon probabilistic genetic information (70), especially when absolute risks remain modest compared to classic lifestyle and environmental factors (9). Furthermore, clinicians may face challenges in interpreting PRS results themselves, which can hinder effective risk communication to patients (71). Second, PRSs still have limited predictive power compared to traditional clinical risk factors (10,72), and this may have reduced their motivational impact. Third, most interventions applied a simple disclosure of PRS results, which may not produce sustained lifestyle change without accompanying substantive behavioral or counseling support (73,74). Evidence from personalized nutrition, similarly suggests that the integration of genetic feedback yields only small and inconsistent effects on dietary change (17,18). For clinicians, these findings highlight that PRS information should not be expected to independently improve adherence or outcomes in preventive care. For policymakers, the results suggest that while PRSs may have some potential to stratify innate risk, they may not have translational impact on behavior change and health outcomes, limiting their clinical value in this setting. Alternatively, PRS may be integrated into predictive algorithms and population stratification strategies (75,76), rather than provided through simple information disclosure. However, there is a need for further research into the integrated use of PRS that evaluates their real world clinical utility in relation to the costs and ethical considerations associated with genomic testing (77).

Several questions remain open. While the evidence to-date is unfavorable for PRS, communication of such information might be more effective in specific populations, such as individuals with high baseline risk, those already motivated to change, or in younger cohorts who have longer lifetime risk horizons (14,78,79). The optimal method for presenting PRS information also requires investigation, including visual risk tools, tailored counseling, and integration with digital health platforms (11). Long-term effects remain largely unexplored, as most trials followed participants for less than one year. Furthermore, most trials were conducted in high-income countries with limited representation of underrepresented groups (80,81). Finally, research on cost-effectiveness (82) and ethical aspects, including equity, privacy, and potential psychological harms (77), will be critical to inform whether PRS disclosure should play a role in any routine preventive medicine program (71). While newer genome-wide PRSs offer improved predictive performance, meeting these additional behavioral, ethical, and translational requirements may remain challenging, thus limiting the real-world utility of personalized medicine tools.

## Ethics statements

### Ethical approval

Not required.

### Contributors

LR: Conceptualization, Methodology, Validation, Formal analysis, Investigation, Data Curation, Writing - Original Draft, Writing - Review & Editing, Visualization. NL: Validation, Formal analysis, Investigation, Data Curation, Writing - Original Draft, Writing - Review & Editing, Visualization. SF: Formal analysis, Investigation, Writing - Original Draft, Writing - Review & Editing. AC, AA, AP: Formal analysis, Investigation, Writing - Review & Editing. RP: Conceptualization, Methodology, Validation, Resources, Writing - Original Draft, Writing - Review & Editing, Supervision CC: Formal analysis, Writing - Review & Editing. SB: Conceptualization, Methodology, Validation, Resources, Writing - Original Draft, Writing - Review & Editing, Supervision, funding acquisition. JPAI: Conceptualization, Methodology, Validation, Formal analysis, Writing - Original Draft, Writing - Review & Editing, Visualization, Supervision. JPAI and SB are the guarantors of the review.

### Data availability statement and code sharing

The full dataset and code used for analysis will be freely available online on the OpenScience Framework website after publication at this link: https://osf.io/28v6j.

### Patient consent for publication

Not applicable.

### Transparency

The lead authors (the manuscript’s guarantors) affirm that the manuscript is an honest, accurate, and transparent account of the study being reported; that no important aspects of the study have been omitted; and that any discrepancies from the study as planned (and, if relevant, registered) have been explained.

### Dissemination to participants and related patient and public communities

We plan to disseminate our findings through various avenues to the public, patients, and clinical organisations, including social media, plain language summaries, and conference presentations. The findings will be communicated in accessible, non-technical language to ensure understanding among general audiences.

## References

1. Manolio TA, Rowley R, Williams MS, Roden D, Ginsburg GS, Bult C, et al. Opportunities, resources, and techniques for implementing genomics in clinical care. The Lancet. 2019 Aug;394(10197):511–20.

2. Farina S, Osti T, Russo L, Maio A, Scarsi N, Savoia C, et al. The current landscape of personalised preventive approaches for non-communicable diseases: A scoping review. PLoS One. 2025 Jan 13;20(1):e0317379.

3. Wellmann R, Borden BA, Danahey K, Nanda R, Polite BN, Stadler WM, et al. Analyzing the clinical actionability of germline pharmacogenomic findings in oncology. Cancer. 2018 Jul 15;124(14):3052–65.

4. Reizine NM, Danahey K, Truong TM, George D, House LK, Karrison TG, et al. Clinically actionable genotypes for anticancer prescribing among >1500 patients with pharmacogenomic testing. Cancer. 2022 Apr 15;128(8):1649–57.

5. Lunke S, Bouffler SE, Patel C V., Sandaradura SA, Wilson M, Pinner J, et al. Integrated multi-omics for rapid rare disease diagnosis on a national scale. Nat Med. 2023 Jul 8;29(7):1681–91.

6. Laurie S, Steyaert W, de Boer E, Polavarapu K, Schuermans N, Sommer AK, et al. Genomic reanalysis of a pan-European rare-disease resource yields new diagnoses. Nat Med. 2025 Jan 17;

7. Bedrosian I, Somerfield MR, Achatz MI, Boughey JC, Curigliano G, Friedman S, et al. Germline Testing in Patients With Breast Cancer: ASCO–Society of Surgical Oncology Guideline. Journal of Clinical Oncology. 2024 Feb 10;42(5):584–604.

8. Sturm AC, Knowles JW, Gidding SS, Ahmad ZS, Ahmed CD, Ballantyne CM, et al. Clinical Genetic Testing for Familial Hypercholesterolemia. J Am Coll Cardiol. 2018 Aug;72(6):662–80.

9. Christoffersen M, Tybjærg-Hansen A. Polygenic risk scores: how much do they add? Curr Opin Lipidol. 2021 Jun;32(3):157– 62.

10. Abu-El-Haija A, Reddi H V., Wand H, Rose NC, Mori M, Qian E, et al. The clinical application of polygenic risk scores: A points to consider statement of the American College of Medical Genetics and Genomics (ACMG). Genetics in Medicine. 2023 May;25(5):100803.

11. Wand H, Lambert SA, Tamburro C, Iacocca MA, O’Sullivan JW, Sillari C, et al. Improving reporting standards for polygenic scores in risk prediction studies. Nature. 2021 Mar 11;591(7849):211–9.

12. Yanes T, McInerney-Leo AM, Law MH, Cummings S. The emerging field of polygenic risk scores and perspective for use in clinical care. Hum Mol Genet. 2020 Oct 20;29(R2):R165–76.

13. Yanes T, McInerney-Leo AM, Law MH, Cummings S. The emerging field of polygenic risk scores and perspective for use in clinical care. Hum Mol Genet. 2020 Oct 20;29(R2):R165–76.

14. Mars N, Koskela JT, Ripatti P, Kiiskinen TTJ, Havulinna AS, Lindbohm J V., et al. Polygenic and clinical risk scores and their impact on age at onset and prediction of cardiometabolic diseases and common cancers. Nat Med. 2020 Apr 7;26(4):549–57.

15. Liyanarachchi S, Gudmundsson J, Ferkingstad E, He H, Jonasson JG, Tragante V, et al. Assessing thyroid cancer risk using polygenic risk scores. Proceedings of the National Academy of Sciences. 2020 Mar 17;117(11):5997–6002.

16. Thomas M, Sakoda LC, Hoffmeister M, Rosenthal EA, Lee JK, van Duijnhoven FJB, et al. Genome-wide Modeling of Polygenic Risk Score in Colorectal Cancer Risk. The American Journal of Human Genetics. 2020 Sep;107(3):432–44.

17. Robinson K, Rozga M, Braakhuis A, Ellis A, Monnard CR, Sinley R, et al. Effect of Incorporating Genetic Testing Results into Nutrition Counseling and Care on Dietary Intake: An Evidence Analysis Center Systematic Review—Part I. J Acad Nutr Diet. 2021 Mar;121(3):553–581.e3.

18. Ellis A, Rozga M, Braakhuis A, Monnard CR, Robinson K, Sinley R, et al. Effect of Incorporating Genetic Testing Results into Nutrition Counseling and Care on Health Outcomes: An Evidence Analysis Center Systematic Review—Part II. J Acad Nutr Diet. 2021 Mar;121(3):582–605.e17.

19. Hollands GJ, French DP, Griffin SJ, Prevost AT, Sutton S, King S, et al. The impact of communicating genetic risks of disease on risk-reducing health behaviour: systematic review with meta-analysis. BMJ. 2016 Mar 15;i1102.

20. Horne J, Madill J, O’Connor C, Shelley J, Gilliland J. A Systematic Review of Genetic Testing and Lifestyle Behaviour Change: Are We Using High-Quality Genetic Interventions and Considering Behaviour Change Theory? Lifestyle Genom. 2018;11(1):49–63.

21. Li SX, Ye Z, Whelan K, Truby H. The effect of communicating the genetic risk of cardiometabolic disorders on motivation and actual engagement in preventative lifestyle modification and clinical outcome: a systematic review and meta-analysis of randomised controlled trials. British Journal of Nutrition. 2016 Sep 14;116(5):924–34.

22. Russo L, Perilli A, Adduci A, Farina S, Pastorino R, Boccia S, et al. 2024. A systematic review and meta-analysis of randomized trials on the efficacy of genetic risk information on behavioral change, therapy adherence and clinical outcomes. doi:10.17605/OSF.IO/28V6J.

23. Page MJ, McKenzie JE, Bossuyt PM, Boutron I, Hoffmann TC, Mulrow CD, et al. The PRISMA 2020 statement: an updated guideline for reporting systematic reviews. BMJ. 2021 May;n71.

24. Babb de Villiers C, Kroese M, Moorthie S. Understanding polygenic models, their development and the potential application of polygenic scores in healthcare. J Med Genet. 2020 Nov;57(11):725–32.

25. Collister JA, Liu X, Clifton L. Calculating Polygenic Risk Scores (PRS) in UK Biobank: A Practical Guide for Epidemiologists. Front Genet. 2022 Feb 18;13.

26. National Cancer Institute. NCI Dictionary of Genetics Terms: PRS. Available at the link: https://www.cancer.gov/publications/dictionaries/genetics-dictionary/def/prs.

27. Dr Sowmiya Moorthie DCB de VTBDLGAHDEJDSS and DMK. Polygenic scores, risk and cardiovascular disease. ISBN978-1-907198-35-9. 2019.

28. D’Ancona F, D’Amario C, Maraglino F, Rezza G, Iannazzo S. The law on compulsory vaccination in Italy: an update 2 years after the introduction. Eurosurveillance. 2019 Jun 27;24(26).

29. Sterne JAC, Savović J, Page MJ, Elbers RG, Blencowe NS, Boutron I, et al. RoB 2: a revised tool for assessing risk of bias in randomised trials. BMJ. 2019 Aug 28;l4898.

30. Hedges LV. Distribution theory for Glass’s estimator of effect size and related estimators. Journal of Educational and Behavioral Statistics. 1981;6:107–28.

31. Luo D, Wan X, Liu J, Tong T. Optimally estimating the sample mean from the sample size, median, mid-range, and/or mid-quartile range. Stat Methods Med Res. 2018 Jun 27;27(6):1785–805.

32. Shi J, Luo D, Weng H, Zeng X, Lin L, Chu H, et al. Optimally estimating the sample standard deviation from the five-number summary. Res Synth Methods. 2020 Sep 25;11(5):641–54.

33. Shi J, Luo D, Wan X, Liu Y, Liu J, Bian Z, et al. Detecting the skewness of data from the five-number summary and its application in meta-analysis. Stat Methods Med Res. 2023 Jul 10;32(7):1338–60.

34. Higgins JPT, Thomas J, Chandler J, Cumpston M, Li T, Page MJ, et al. Cochrane. 2024. Cochrane Handbook for Systematic Reviews of Interventions version 6.5 (updated August 2024). Available from www.training.cochrane.org/handbook.

35. Viechtbauer W. Bias and Efficiency of Meta-Analytic Variance Estimators in the Random-Effects Model. Journal of Educational and Behavioral Statistics. 2005 Sep 1;30(3):261–93.

36. Woodward M. Epidemiology: Study Design and Data Analysis. Chapman and Hall/CRC; 2013.

37. Halmesvaara O, Vornanen M, Kääriäinen H, Perola M, Kristiansson K, Konttinen H. Psychosocial Effects of Receiving Genome-Wide Polygenic Risk Information Concerning Type 2 Diabetes and Coronary Heart Disease: A Randomized Controlled Trial. Front Genet. 2022 May 30;13.

38. McVay MA, Beadles C, Wu R, Grubber J, Coffman CJ, Yancy WS, et al. Effects of provision of type 2 diabetes genetic risk feedback on patient perceptions of diabetes control and diet and physical activity self-efficacy. Patient Educ Couns. 2015 Dec;98(12):1600–7.

39. Livingstone KM, Celis-Morales C, Navas-Carretero S, San-Cristobal R, Macready AL, Fallaize R, et al. Effect of an Internet-based, personalized nutrition randomized trial on dietary changes associated with the Mediterranean diet: the Food4Me Study. Am J Clin Nutr. 2016 Aug;104(2):288–97.

40. Celis-Morales C, Livingstone KM, Marsaux CFM, Macready AL, Fallaize R, O’Donovan CB, et al. Effect of personalized nutrition on health-related behaviour change: evidence from the Food4me European randomized controlled trial. Int J Epidemiol. 2016 Aug 14;dyw186.

41. Vassy JL, He W, Florez JC, Meigs JB, Grant RW. Six-Year Diabetes Incidence After Genetic Risk Testing and Counseling: A Randomized Clinical Trial. Diabetes Care. 2018 Mar 1;41(3):e25–6.

42. Raghavan S, Xu K, Coffman CJ, Pabich S, Edelman D, Voils CI. Associations of Diabetes Genetic Risk Counseling with Incident Diabetes and Weight: 5-Year Follow-up of a Randomized Controlled Trial. J Gen Intern Med. 2020 Mar 16;35(3):944– 6.

43. Naderian M, Hamed ME, Vaseem AA, Norland K, Dikilitas O, Teymourzadeh A, et al. Effect of Disclosing a Polygenic Risk Score for Coronary Heart Disease on Adverse Cardiovascular Events. Circ Genom Precis Med. 2025 Apr;18(2).

44. Nielsen DE, El-Sohemy A. Disclosure of Genetic Information and Change in Dietary Intake: A Randomized Controlled Trial. PLoS One. 2014 Nov 14;9(11):e112665.

45. Weinberg DS, Myers RE, Keenan E, Ruth K, Sifri R, Ziring B, et al. Genetic and Environmental Risk Assessment and Colorectal Cancer Screening in an Average-Risk Population. Ann Intern Med. 2014 Oct 21;161(8):537–45.

46. Hietaranta-Luoma HL, Tahvonen R, Iso-Touru T, Puolijoki H, Hopia A. An Intervention Study of Individual, apoE Genotype-Based Dietary and Physical-Activity Advice: Impact on Health Behavior. Lifestyle Genom. 2014;7(3):161–74.

47. Voils CI, Coffman CJ, Grubber JM, Edelman D, Sadeghpour A, Maciejewski ML, et al. Does Type 2 Diabetes Genetic Testing and Counseling Reduce Modifiable Risk Factors? A Randomized Controlled Trial of Veterans. J Gen Intern Med. 2015 Nov 16;30(11):1591–8.

48. Doménech M, Elosua R, Salas E, Sierra C, Marrugat J, Coca A. Awareness of Genetic Coronary Risk Score Improves Blood Pressure Control in Hypertensive Patients. Revista Española de Cardiología (English Edition). 2016 Dec;69(12):1226–7.

49. Wu RR, Myers RA, Hauser ER, Vorderstrasse A, Cho A, Ginsburg GS, et al. Impact of Genetic Testing and Family Health History Based Risk Counseling on Behavior Change and Cognitive Precursors for Type 2 Diabetes. J Genet Couns. 2017 Feb 14;26(1):133–40.

50. Horne JR, Gilliland J, Leckie T, O’Connor C, Seabrook JA, Madill J. Can a Lifestyle Genomics Intervention Motivate Patients to Engage in Greater Physical Activity than a Population-Based Intervention? Results from the NOW Randomized Controlled Trial. Lifestyle Genom. 2020;13(6):180–6.

51. Lacson JCA, Doyle SH, Qian L, Del Rio J, Forgas SM, Valavanis S, et al. A Randomized Trial of Precision Prevention Materials to Improve Primary and Secondary Melanoma Prevention Activities among Individuals with Limited Melanoma Risk Phenotypes. Cancers (Basel). 2021 Jun 23;13(13):3143.

52. Watanabe M, Hosono S, Nakagawa-Senda H, Yamamoto S, Aoyama M, Hattori S, et al. Does Direct-to-Consumer Personal Genetic Testing Improve Gynecological Cancer Screening Uptake among Never-Screened Attendees? A Randomized Controlled Study. Int J Environ Res Public Health. 2021 Nov 24;18(23):12333.

53. Lacson JCA, Doyle SH, Del Rio J, Forgas SM, Carvajal R, Gonzalez-Calderon G, et al. A Randomized Clinical Trial of Precision Prevention Materials Incorporating *MC1R* Genetic Risk to Improve Skin Cancer Prevention Activities Among Hispanics. Cancer Research Communications. 2022 Jan 11;2(1):28–38.

54. Wolever RQ, Yang Q, Maldonado CJ, Armitage NH, Musty MD, Kraus WE, et al. Health coaching and genetic risk testing in primary care: Randomized controlled trial. Health Psychology. 2022 Oct;41(10):719–32.

55. Wu YP, Hamilton JG, Kaphingst KA, Jensen JD, Kohlmann W, Parsons BG, et al. Increasing Skin Cancer Prevention in Young Adults: the Cumulative Impact of Personalized UV Photography and MC1R Genetic Testing. Journal of Cancer Education. 2023 Jun 28;38(3):1059–65.

56. Halmesvaara O, Lonna M, Kääriäinen H, Perola M, Kristiansson K, Konttinen H. The impact of supplementing traditional risk information with polygenic risk score concerning type 2 diabetes and coronary heart disease on health behavior: a randomized controlled trial. J Community Genet. 2025 Mar 26;

57. Knowles JW, Zarafshar S, Pavlovic A, Goldstein BA, Tsai S, Li J, et al. Impact of a Genetic Risk Score for Coronary Artery Disease on Reducing Cardiovascular Risk: A Pilot Randomized Controlled Study. Front Cardiovasc Med. 2017 Aug 14;4.

58. Owaki Y, Yoshimoto H, Saito G, Dobashi S, Kushio S, Nakamura A, et al. Effectiveness of genetic feedback on alcohol metabolism to reduce alcohol consumption in young adults: an open-label randomized controlled trial. BMC Med. 2024 May 20;22(1):205.

59. Lee GY, Chung KM, Lee J, Kim JH, Han SN. Changes in anxiety and depression levels and meat intake following recognition of low genetic risk for high body mass index, triglycerides, and lipoproteins: A randomized controlled trial. PLoS One. 2023 Sep 8;18(9):e0291052.

60. Viigimaa M, Jürisson M, Pisarev H, Kalda R, Alavere H, Irs A, et al. Effectiveness and feasibility of cardiovascular disease personalized prevention on high polygenic risk score subjects: a randomized controlled pilot study. European Heart Journal Open. 2022 Nov 1;2(6).

61. Ma RCW, Xie F, Lim CKP, Lau ESH, Luk AOY, Ozaki R, et al. A randomized clinical trial of genetic testing and personalized risk counselling in patients with type 2 diabetes receiving integrated care -The genetic testing and patient empowerment (GEM) trial. Diabetes Res Clin Pract. 2022 Jul;189:109969.

62. Smit AK, Espinoza D, Newson AJ, Morton RL, Fenton G, Freeman L, et al. A Pilot Randomized Controlled Trial of the Feasibility, Acceptability, and Impact of Giving Information on Personalized Genomic Risk of Melanoma to the Public. Cancer Epidemiology, Biomarkers & Prevention. 2017 Feb 1;26(2):212–21.

63. Silarova B, Sharp S, Usher-Smith JA, Lucas J, Payne RA, Shefer G, et al. Effect of communicating phenotypic and genetic risk of coronary heart disease alongside web-based lifestyle advice: The INFORM Randomised Controlled Trial. Heart. 2019 Jul 1;105(13):982–9.

64. Smit AK, Allen M, Beswick B, Butow P, Dawkins H, Dobbinson SJ, et al. Impact of personal genomic risk information on melanoma prevention behaviors and psychological outcomes: a randomized controlled trial. Genetics in Medicine. 2021 Dec;23(12):2394–403.

65. Nichols JAA, Grob P, Kite W, Williams P, de Lusignan S. Using a genetic/clinical risk score to stop smoking (GeTSS): randomised controlled trial. BMC Res Notes. 2017 Dec 23;10(1):507.

66. Kullo IJ, Jouni H, Austin EE, Brown SA, Kruisselbrink TM, Isseh IN, et al. Incorporating a genetic risk score into coronary heart disease risk estimates: Effect on low-density lipoprotein cholesterol levels (the MI-GENES Clinical Trial). Circulation. 2016 Mar 22;133(12):1181–8.

67. Turner AR, Lane BR, Rogers D, Lipkus I, Weaver K, Danhauer SC, et al. Randomized trial finds that prostate cancer genetic risk score feedback targets prostate-specific antigen screening among at-risk men. Cancer. 2016 Nov 15;122(22):3564–75.

68. Godino JG, van Sluijs EMF, Marteau TM, Sutton S, Sharp SJ, Griffin SJ. Lifestyle Advice Combined with Personalized Estimates of Genetic or Phenotypic Risk of Type 2 Diabetes, and Objectively Measured Physical Activity: A Randomized Controlled Trial. PLoS Med. 2016 Nov 29;13(11):e1002185.

69. Grant RW, O’Brien KE, Waxler JL, Vassy JL, Delahanty LM, Bissett LG, et al. Personalized Genetic Risk Counseling to Motivate Diabetes Prevention. Diabetes Care. 2013 Jan 1;36(1):13–9.

70. Lewis ACF, Perez EF, Prince AER, Flaxman HR, Gomez L, Brockman DG, et al. Patient and provider perspectives on polygenic risk scores: implications for clinical reporting and utilization. Genome Med. 2022 Oct 7;14(1):114.

71. Andreoli L, Peeters H, Van Steen K, Dierickx K. Polygenic risk scores in the clinic: a systematic review of stakeholders’ perspectives, attitudes, and experiences. European Journal of Human Genetics. 2025 Mar 23;33(3):266–80.

72. Groenendyk JW, Greenland P, Khan SS. Incremental Value of Polygenic Risk Scores in Primary Prevention of Coronary Heart Disease. JAMA Intern Med. 2022 Oct 1;182(10):1082.

73. Wallingford CK, Kovilpillai H, Jacobs C, Turbitt E, Primiero CA, Young MA, et al. Models of communication for polygenic scores and associated psychosocial and behavioral effects on recipients: A systematic review. Genetics in Medicine. 2023 Jan;25(1):1–11.

74. Widén E, Junna N, Ruotsalainen S, Surakka I, Mars N, Ripatti P, et al. How Communicating Polygenic and Clinical Risk for Atherosclerotic Cardiovascular Disease Impacts Health Behavior: an Observational Follow-up Study. Circ Genom Precis Med. 2022 Apr;15(2).

75. Mosley JD, Gupta DK, Tan J, Yao J, Wells QS, Shaffer CM, et al. Predictive Accuracy of a Polygenic Risk Score Compared With a Clinical Risk Score for Incident Coronary Heart Disease. JAMA. 2020 Feb 18;323(7):627.

76. Elliott J, Bodinier B, Bond TA, Chadeau-Hyam M, Evangelou E, Moons KGM, et al. Predictive Accuracy of a Polygenic Risk Score–Enhanced Prediction Model vs a Clinical Risk Score for Coronary Artery Disease. JAMA. 2020 Feb 18;323(7):636.

77. Sanderson SC, Inouye M. Psychological and behavioural considerations for integrating polygenic risk scores for disease into clinical practice. Nat Hum Behav. 2025 May 12;9(6):1098–106.

78. Ratman D, Tshiaba P, Levin M, Sun J, Tunstall T, Maier R, et al. Polygenic risk scores improve CAD risk prediction in individuals at borderline and intermediate clinical risk. npj Cardiovascular Health. 2025 May 1;2(1):13.

79. Samani NJ, Beeston E, Greengrass C, Riveros-McKay F, Debiec R, Lawday D, et al. Polygenic risk score adds to a clinical risk score in the prediction of cardiovascular disease in a clinical setting. Eur Heart J. 2024 Sep 7;45(34):3152–60.

80. Kullo IJ, Conomos MP, Nelson SC, Adebamowo SN, Choudhury A, Conti D, et al. The PRIMED Consortium: Reducing disparities in polygenic risk assessment. The American Journal of Human Genetics. 2024 Dec;111(12):2594–606.

81. Kullo IJ. Promoting equity in polygenic risk assessment through global collaboration. Nat Genet. 2024 Sep 5;56(9):1780–7.

82. Siena LM, Baccolini V, Riccio M, Rosso A, Migliara G, Sciurti A, et al. Weighing the evidence on costs and benefits of polygenic risk-based approaches in clinical practice: A systematic review of economic evaluations. The American Journal of Human Genetics. 2025 Aug;112(8):1735–53.

